# Drug-drug interactions between classic psychedelics and psychoactive drugs: a systematic review

**DOI:** 10.1101/2023.06.01.23290811

**Authors:** Andreas Halman, Geraldine Kong, Jerome Sarris, Daniel Perkins

## Abstract

Classic psychedelics, lysergic acid diethylamide, psilocybin, mescaline and N,N-dimethyltryptamine, are potent psychoactive substances that have been studied for their physiological and psychological effects. However, our understanding of the potential interactions and outcomes of using these substances are used in combination with other psychoactive drugs is limited.

This systematic review aims to provide a comprehensive overview of the current research on drug-drug interactions between classic psychedelics and other psychoactive drugs in humans. We conducted a thorough literature search using multiple databases, including PubMed, PsycINFO, Web of Science and other sources to supplement our search for relevant studies. A total of 8,487 records were screened, and studies involving human data describing potential interactions (as well as the lack thereof) between classic psychedelics and other psychoactive drugs were included.

In total, we identified 50 studies from 34 reports published before April 20, 2023, encompassing 31 studies on LSD, 11 on psilocybin, 4 on mescaline, 3 on DMT and 1 on ayahuasca. These studies provide insights into the interactions between classic psychedelics and a range of drugs, including antidepressants, antipsychotics, anxiolytics, mood stabilisers, recreational drugs and others.

The findings revealed various effects when psychedelics were combined with other drugs, including both attenuated and potentiated effects, as well as instances where no changes were observed. Except for a few case reports, no serious adverse drug events were described in the included studies. In-depth discussion of the results is presented, along with an exploration of the potential molecular pathways that underlie the observed effects.

## Introduction

Classic psychedelics, including lysergic acid diethylamide (LSD), psilocybin (psilocin as an active agent), mescaline and N,N-dimethyltryptamine (DMT), can be structurally divided into two classes: tryptamines (psilocybin/psilocin, DMT and LSD) and phenethylamines (mescaline) (Johnson et al., 2008). While LSD was first synthesised in 1938, mescaline, psilocybin and DMT are all naturally occurring substances that have been used by humans for centuries (Schlag et al., 2022).

**LSD** served as an experimental tool during the 1950s-1970s for studying psychotic-like states and modelling psychosis (Bercel et al., 1956), as well as being used in psycholytic psychotherapy (Grieco & Bloom, 1981). Potential therapeutic applications of LSD have also been explored, including its use in treating alcoholism (Smart et al., 1966), addiction (Savage & McCabe, 1973) and terminal cancer (Grof et al., 1973). However, the widespread unregulated use of LSD led to the cessation of clinical research in the early 1970s due to political pressure (Liechti, 2017). Despite this, LSD research, along with other classic psychedelics, is picking up speed and emerging again as a powerful tool for treating psychiatric disorders (Carhart-Harris et al., 2021; Davis et al., 2021; Gasser et al., 2015; Holze et al., 2023; Schindler et al., 2021; von Rotz et al., 2023).

The average dose of LSD is 50-200 µg (Dinis-Oliveira et al., 2019) and the effects of a moderate dose (100 µg) last on average for around 8 hours (Ley et al., 2023). The subjective effects typically involve visual hallucinations, positive experiences of derealization and depersonalization, audiovisual synaesthesia, and potential increases in feelings of well-being, happiness, closeness to others, openness and trust (Schmid et al., 2015). Physically, it can increase blood pressure, heart rate, body temperature, pupil dilation, plasma cortisol, prolactin, oxytocin and epinephrine (Schmid et al., 2015). However, LSD, as well as other psychedelics, can also have adverse effects and users may experience “bad trips” characterised by anxiety, paranoia, fear and dysphoria (Johnson et al., 2008). In rare cases, the use of LSD can trigger the onset of prolonged psychosis or cause hallucinogen persisting perception disorder (Johnson et al., 2008). LSD has a high affinity for several serotonin (5-HT) receptors (such as 5-HT_1A/B_ and 5-HT_2A_) and has affinity to dopaminergic D_1-5_ receptors (Halberstadt & Geyer, 2011). The affinity for 5-HT_2A_ is important as the main mechanism behind the behavioural and psychological effects of LSD and other psychedelics are thought to be the activation of 5-HT_2A_ receptors in cortical and subcortical structures (Vollenweider & Smallridge, 2022). LSD is metabolised by enzymes such as CYP1A2, CYP3A4, CYP2C9, CYP2C19 and CYP2D6, with particular emphasis on the primary contribution of the former two (Wagmann et al., 2019).

While LSD is chemically synthesised, **psilocybin** is a naturally occurring psychedelic compound found in various genera of mushrooms, including *Psilocybe, Panaeolus*, *Conocybe*, *Gymnopilus, Stropharia, Pluteus and Panaeolina* (Dinis-Oliveira et al., 2019). These mushrooms are commonly referred to as “magic mushrooms’’ due to their psychedelic and hallucinogenic effects (Dinis-Oliveira, 2017). The average dose is 20-40 mg (Dinis-Oliveira et al., 2019) and a moderate dose of 20 mg typically results in effects lasting around 5 hours (Ley et al., 2023). Typically, the effects involve changes in perception, cognition, affect and mystical-type experiences (Griffiths et al., 2011), similar in nature to those experienced with LSD. Once ingested, the body metabolises psilocybin to psilocin, which is the primary psychoactive compound (Dinis-Oliveira, 2017). The metabolism of psilocybin involves several enzymes, including aldehyde dehydrogenase (ALDH), monoamine oxidase (MAO), UGT1A9, UGT1A10, UGT1A6, UGT1A7 and UGT1A8 enzymes (Dinis-Oliveira, 2017). Similar to LSD, psilocin acts as an agonist at 5-HT_2A_ receptors in the brain (Rickli et al., 2016), where it exerts its psychological effects (Vollenweider et al., 1998).

**Mescaline** (3,4,5-trimethoxyphenethylamine) is a psychedelic compound found in the North American peyote cactus (*Lophophora williamsii*), the South American San Pedro cactus (*Echinopsis pachanoi*), as well as other cacti such as the Peruvian torch cactus (*Echinopsis peruviana*), Bolivian torch cactus (*Echinopsis lageniformis*) and the leaf cactus (*Pereskia aculeata*) (Dinis-Oliveira et al., 2019). It has high affinity for 5-HT_1A_ and 5-HT_2A/C_ receptors, but is less potent than LSD, psilocin and DMT (Rickli et al., 2016). Mescaline is metabolised in the liver and broken down into several inactive compounds, with oxidative deamination occurring via MAO or diamine oxidase (DAO) (Dinis-Oliveira et al., 2019). Mescaline is approximately 30 times less potent than psilocybin, and a moderate dose corresponds to 200-400 mg of mescaline sulphate (Ley et al., 2023). The effects of mescaline last longer than those of LSD, with an average duration of about 11 hours and peak effects occurring around 4 hours after administration (Ley et al., 2023). The subjective effects of mescaline are similar to those of other classic psychedelics and may include hallucinations (closed and open-eye visuals, synaesthesia), altered thinking processes, altered sense of time and spatial distortion (Dinis-Oliveira et al., 2019).

Finally, **N,N-dimethyltryptamine (DMT)** is a naturally occurring psychoactive compound found in several plants and is also endogenously produced in mammals, including humans (Jiménez & Bouso, 2022). Recreationally, DMT is consumed either in a pure form (mostly smoked) or as a key ingredient in an orally active brew called ayahuasca (Cakic et al., 2010). Ayahuasca is made by mixing a DMT-containing plant with a vine containing β-carboline alkaloids (Brito-da-Costa et al., 2020). Although the exact plant combinations can vary, a frequent mixture is made of *Psychotria viridis* (source of DMT) and *Banisteriopsis caapi* vine, which stem and bark contains β-carbolines harmine and harmaline (Brito-da-Costa et al., 2020).

DMT is a partial agonist primarily of the 5-HT_1A_, 5-HT_2A_ and 5-HT_2C_ receptors (Carbonaro & Gatch, 2016). The effects of DMT can vary depending on the route of administration and dosage. When inhaled or injected, the onset of effects is rapid (Barker, 2022). After injection, the effects peak around 90-120 seconds and last up to 30 minutes, during which users may experience intense visual hallucinations, euphoria and anxiety (Strassman et al., 1994). Oral consumption of DMT does not produce psychotropic effects due to rapid metabolism by MAO enzymes (Riba et al., 2015). However, when consumed orally as part of the ayahuasca brew, DMT becomes bioavailable due to the MAO-A inhibiting effects of harmine and harmaline, which protect DMT from the deamination in the gut (Brito-da-Costa et al., 2020). In this case, the effects are typically felt within 30-60 minutes and last for around four hours (Barker, 2022). DMT itself has a short half-life of 9-12 minutes and is rapidly metabolised by MAO-A, while CYP2D6 and, to a lesser extent, CYP2C19 are also involved in its metabolism (Good et al., 2022).

**Drug-drug interactions (DDIs)** can be categorised as either pharmacokinetic or pharmacodynamic interactions. Pharmacokinetic interactions occur when one drug influences the absorption, distribution, metabolism or elimination of another drug. On the other hand, pharmacodynamic interactions involve the modification of the pharmacological effect of one drug by another. These interactions can exhibit synergistic, additive or antagonistic characteristics. Additivity refers to the overall effect of a drug combination which is the sum of the effects of each individual drug, while synergy occurs when the combined effect of the drugs is greater than additive. Antagonism arises when the combined effect is less than additive (Niu et al., 2019).

One common mechanism of pharmacodynamic drug interaction is competition at the receptor level. When two drugs interact with the same receptor, they can compete for binding, leading to alterations in their pharmacological effects (Lambert, 2004). For instance, blocking the receptors where LSD, psilocin, mescaline or DMT exert their effects, such as 5-HT_2A_, could impede their psychological effects.

An example of a pharmacokinetic DDI is the inhibition of drug-metabolising enzymes, such as cytochrome P450 (CYP) that are responsible for metabolising a broad range of drugs (Zhao et al., 2021). Inhibition of these enzymes by concomitant drugs or circulating metabolites can lead to altered drug metabolism and impact the drug’s effects and influence treatment outcomes (Zhao et al., 2021). Additionally, there is the potential for interaction with P-glycoprotein (P-gp), a membrane transporter that facilitates the efflux of various drugs and is present in kidneys, liver, gastrointestinal tract and blood-brain barrier (M. L. Amin, 2013). Similar to CYP enzymes, reducing the activity of P-gp can increase the concentration of its substrates in the blood, whereas increasing its activity can decrease the concentration, leading to inadequate therapeutic effects (M. L. Amin, 2013).

For instance, CYP enzymes have a known role in LSD metabolism (Luethi et al., 2019), and therefore can affect LSD’s effects. DDIs can occur even when drugs are not taken concurrently, allowing for days or even weeks between their administration. Some drugs, like fluoxetine, have prolonged inhibitory effects on CYP-activity that may persist for several weeks following its discontinuation due to the extended half-life of fluoxetine and its metabolite norfluoxetine (Hemeryck & Belpaire, 2002).

Currently, there is limited literature available on the drug-drug interactions between classic psychedelics and other psychoactive drugs. While two review articles have been published on this subject, the first one was limited in scope (Wyatt et al., 1976) and the second one focused solely on MDMA and psilocybin interactions with psychiatric medications (Sarparast et al., 2022). To fill this gap, this systematic review aimed to provide a comprehensive overview of the current state of research on DDIs between classic psychedelics and any other psychoactive drugs. We conducted a thorough literature search and reported on both physiological and subjective outcomes. This review offers valuable insights into the potential risks and benefits of combining classic psychedelics with other psychoactive drugs, thereby guiding researchers and clinicians in this field.

## Methods

This review was registered in PROSPERO (CRD42022336092) and followed the latest PRISMA (2020) guidelines (Page et al., 2021). A keyword search for articles pertaining to classical psychedelics (LSD, psilocybin, mescaline, DMT and ayahuasca) was initially conducted on June 5, 2022, in three primary scientific databases: PubMed, PsycINFO and Web of Science (with no year restriction). These databases comprise a general journal articles database and a database specialised in biomedicine and psychology research. Search terms included keywords (including synonyms) related to classic psychedelics in the title, abstract, keywords, full text (where available) and MeSH terms, as well as keywords related to drug interactions, side-effects and adverse reactions. The search was not limited by time period in order to capture all relevant articles. A total of 2,151 articles were found during the first search. The search was repeated on April 20, 2023, to manually identify any additional articles published since the initial search. The full search terms for each database can be found in the Supplementary information.

Additionally, to ensure comprehensive coverage, a search was conducted in the Multidisciplinary Association for Psychedelic Studies (MAPS) comprehensive online “Psychedelic Bibliography” database (https://bibliography.maps.org), which contains scientific but also non-scientific articles specifically about psychedelics, dating back to 1841. At the time of the search (20 Nov 2022), the database included a total of 13,237 records. All records were downloaded, followed by keyword search in all titles and abstracts (targeting “LSD”, “psilocybin”, “mescaline”, “DMT” and “ayahuasca”, including synonyms and variations), which resulted in a total of 6,336 records.

Furthermore, we manually conducted a search in a registry of clinical trials at ClinicalTrials.gov on April 20, 2023, to find any articles containing results from clinical trials that were missed in the database search and conducted reference lists checking of included studies to find additional missing articles.

Records were screened in two phases: firstly, the ones found via scientific database search and after the completion, the ones found in the MAPS bibliography database. In both cases, a systematic review application Catchii (Halman & Oshlack, 2023) was used. Duplicate removal was done by using Catchii’s duplicate detection and manually verifying the results before removal. Two researchers independently conducted title and abstract screening, both being blind to each other’s decisions. The inclusion criteria were: (A) any article on human participants, (B) any article describing usage of any classical psychedelics with another psychoactive drug, (C) studies describing subjective and/or psychological effects of classical psychedelics with another psychoactive drug (or the lack of), (D) studies that were either randomised controlled trials, observational studies (cohort, case-control, cross-sectional) or case reports/studies, (E) studies published in English. Disagreements between reviewers were discussed until a consensus decision was reached. Records that did not meet the population, intervention and outcome (PIO) criteria were excluded. A total of 8,487 records were screened in the first stage (title and abstract analysis) and full-text of 75 reports were assessed in the second stage (including articles identified from citation search). The summary of results can be seen on the PRISMA flow diagram (Figure 1). Data from all eligible records was extracted by authors of this review, who corroborated each other’s findings. Outcome measures included physiological and subjective effects.

**Figure 1.**
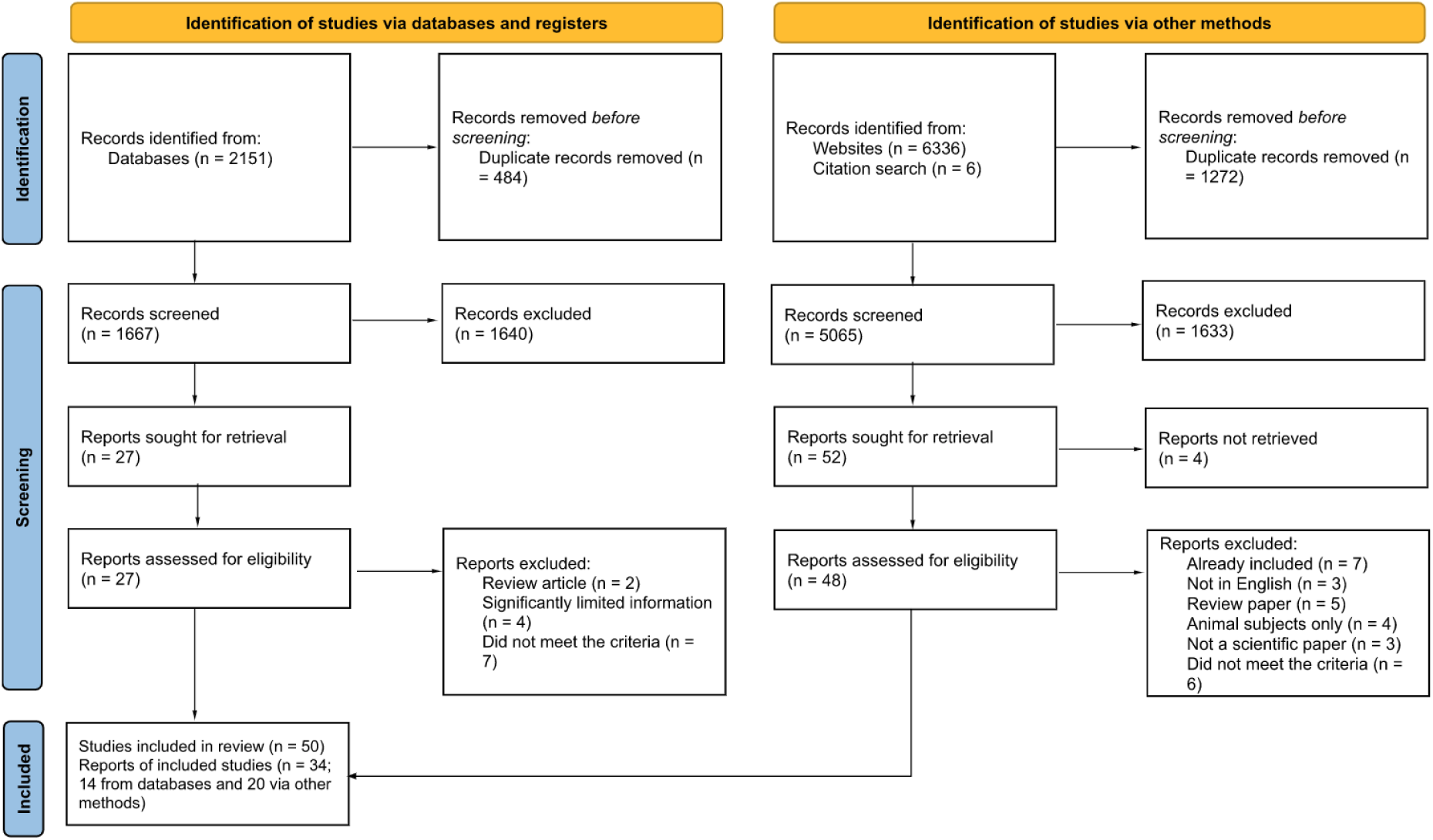
PRISMA flow diagram.

## Results

The results of this systematic review include studies investigating drug-drug interactions between psychedelic drugs (LSD, psilocybin, mescaline, DMT and ayahuasca) and other psychoactive drugs. Regarding LSD, the review examined interactions with antipsychotics (chlorpromazine), mood stabilisers (lithium), various antidepressants including selective serotonin reuptake inhibitors (SSRIs; fluoxetine, sertraline, paroxetine and trazodone), tricyclic antidepressants (TCAs; imipramine, desipramine and clomipramine), as well as monoamine oxidase inhibitors (MAOIs; phenelzine, isocarboxazid, nialamide and iproniazid) and other substances (azacyclonol). Recreational substances (alcohol), as well as other substances such as ketanserin, reserpine, niacin, scopolamine and phenoxybenzamine, were also included.

Regarding psilocybin, the review contains reports on its interactions with anxiolytics (buspirone), antipsychotics (chlorpromazine, haloperidol and risperidone), SSRI antidepressants (escitalopram) and recreational drugs (alcohol and cannabis). Other investigated substances included ketanserin and ergotamine. In the case of mescaline, the review examined interactions with antipsychotics (chlorpromazine and promazine), antidepressants (azacyclonol) and the compound 2,4,5-trimethoxyphenethylamine (2C-O). For DMT, the review explored interactions with MAOI antidepressants (iproniazid), as well as other substances like racemic pindolol and methysergide. Lastly, one report of the SSRI antidepressant fluoxetine was found for ayahuasca.

Separate tables are provided in this review for LSD (Table 1), psilocybin (Table 2), mescaline (Table 3) and DMT together with ayahuasca (Table 4), with results sorted by drug class. Additionally, a combined Table 5 summarises all results from Tables 1-4, along with the main molecular targets of each drug.

**Table 1.** Summary of studies and case reports describing interactions (or the absence thereof) between LSD and other psychoactive drugs.

| Author and year | Study type | Drug | Dose | Participants | Outcome |
| --- | --- | --- | --- | --- | --- |
| (Isbell & Logan, 1957) | Placebo-controlled trial | Antipsychotics:<br>Chlorpromazine | 1) Pretreatment with 50-100 mg chlorpromazine 30 mins prior to 40-60 µg LSD.<br>2) Administration of chlorpromazine at the height of LSD reaction.<br>3) Administration of either 75 mg oral or 25 mg intramuscular chlorpromazine 30 mins after ingestion of 60-150 µg LSD | N = 6-19 (adult male drug addicts) | Pretreatment of chlorpromazine significantly reduced mental, hallucinations, visual perception distortion, anxiety and pupillary reactions of LSD.<br>Intramuscular, but not oral, administration of chlorpromazine after ingestion of LSD significantly blocked the effects of LSD. |
| (Murphree, 1962) | Controlled trial | Antipsychotics:<br>Chlorpromazine | Pretreatment with 25 mg chlorpromazine 30 mins before ingestion of 15-20 µg LSD.<br>Co-administration of chlorpromazine with LSD. | N = 18 (healthy) | Pretreatment of chlorpromazine 30 mins before LSD blocked LSD effects and elevated the threshold dose. When chlorpromazine was given 30 simultaneously or after LSD ingestion, no blocking effects were observed. |
| (Abramson et al., 1960) | Controlled trial | Antipsychotics:<br>Chlorpromazine | Three experiments of oral administration of 50 mg chlorpromazine at 1.5 hours before, after, or simultaneous with 50 µg of LSD. | N = 3 (healthy) | Participants who took chlorpromazine before LSD ingestion showed enhanced positive responses of LSD. Chlorpromazine ingestion after LSD also increased positive responses of LSD and reduced the usual anxiety symptoms such as sweating and moist palms. Simultaneous ingestion of chlorpromazine and LSD produced similar effects as LSD alone. |
| (Schwarz, 1967) | Case series | Antipsychotics:<br>Chlorpromazine | 25 mg chlorpromazine was given 4 hours after 100 µg of LSD (intravenous). | N = 2 (schizophrenic)<br>Age: 22 and 33 | Both patients reported that chlorpromazine administration increased anxiety and intensified effects of LSD. |
| (Bonson & Murphy, 1996) | Case series | Mood stabilisers:<br>Lithium | 600-1000 mg/day of lithium for 7-50 weeks prior to 200 µg of LSD. | N = 3 (patients treated for depression)<br>Age: 21-29 | Participants experienced earlier onset as well as increased hallucinatory and psychological effects of LSD. |
| (Strassman, 1992) | Case report | Antidepressants (SSRI):<br>Fluoxetine | 20 mg/day fluoxetine; LSD dose unknown but one that produces full hallucinogenic effects. | N = 1 (patient for dysthymia)<br>Age: 38 | Markedly decreased in sensitivity to LSD and required 2 to 3 times more than the usual dose to elicit a full hallucinogenic effect of LSD. |
| (Bonson et al., 1996) | Cross-sectional study | Antidepressants (SSRI): Fluoxetine | Pretreatment of fluoxetine of 20-80 mg/day for 2-150 weeks followed by 250 µg of LSD. | N = 18 (patients treated for depression) | Fluoxetine delayed the onset of LSD effects (8 out of 18) and markedly diminished the hallucinogenic and psychological effects as well as overall response of LSD regardless of dose or duration of fluoxetine pretreatment. |
| (Bonson et al., 1996) | Cross-sectional study | Antidepressants (SSRI): Sertraline | Pretreatment of sertraline of 50-200 mg/day for 3-40 weeks followed by 200 µg of LSD. | N = 11 (patients treated for depression) | Sertraline did not affect the onset of LSD effects. Most of the participants experienced decreased physical, hallucinogenic and psychological effects of LSD regardless of dosage or duration of sertraline treatment. |
| (Bonson et al., 1996) | Cross-sectional study | Antidepressants (SSRI): Paroxetine | Pretreatment of 20-40 mg/day paroxetine for 3-30 weeks followed by 150 µg of LSD. | N = 3 (patients treated for depression) | Paroxetine pretreatment did not affect onset of effects but attenuated hallucinogenic as well as physiologic effects of LSD and overall, reduced responses of LSD. |
| (Bonson et al., 1996) | Case report | Antidepressants (SARI): Trazodone | Pretreatment with 200 mg/day for 24 weeks followed by 'moderate' dose of LSD (no more details given). | N = 1 (patient treated for depression) | No changes to the onset of LSD effects but reduced the hallucinogenic and physiological effects of LSD as well as overall response of LSD. |
| (Bonson & Murphy, 1996) | Case series | Antidepressants (TCA): Imipramine | 175-200 mg/day for 8-40 weeks before LSD dose of 80-200 µg. | N = 2 (patients treated for depression)<br>Age: 26-28 | Patients experienced earlier onset of LSD effects and increased hallucinatory as well as psychological effects. |
| (Bonson & Murphy, 1996) | Case series | Antidepressants (TCA): Desipramine | 200 mg/day for 100-150+ weeks before 100-150 µg of LSD dose. | N = 2 (patients treated for depression)<br>Age: 27-32 | Patients experienced earlier onset of LSD effects and increased hallucinatory as well as psychological effects. |
| (Bonson & Murphy, 1996) | Case report | Antidepressants (TCA): Clomipramine | 125 mg/day for 12 weeks before a 'moderate' dose of LSD. | N = 1 (patient treated for alcoholism)<br>Age: 25 | Patient experienced earlier onset of LSD effect, increased physical and hallucinatory as well as psychological effects. |
| (Bonson & Murphy, 1996) | Case series | Antidepressants (MAOI): Phenelzine | 60-75 mg/day for 12 weeks before 150 µg of LSD. | N = 2 (patients treated for depression) | No change to the onset of LSD effects but nearly abolished the subjective responses of LSD including hallucination and psychological effects |
| (Resnick et al., 1964) | Controlled trial | Antidepressants (MAOI): Isocarboxazid | Pretreatment with 30 mg/day isocarboxazid for 5 days or 2 weeks before 40 µg of oral ingestion of | N = 4 (healthy) | Isocarboxazid pretreatment attenuated psychological, autonomic and neurologic responses of LSD including blood pressure, heart rate, sensory and motor functions. |
|  |  |  | LSD-25. |  |  |
| (Grof & Dytrych, 1965) | Controlled trial | Antidepressants (MAOI): Nialamide | 250-500 mg/day of parenteral administration of nialamide for several days, followed by 150-300 mg/day peroral application of nialamide, and then 150 µg of LSD on test day.<br>Pretreatment of nialamide daily for 3 weeks to reach a total dose of 3500 mg, followed by 150 µg of LSD on test day. | N = 14 (neurotic patients) | Premedication with nialamide blocked reactions to LSD and patients were able to tolerate 500 µg of LSD without clinical or psychotic reactions/symptoms of LSD and minimal subjective complaints. |
| (DeMaar et al., 1960) | Placebo-controlled trial | Antidepressants (MAOI): Iproniazid | 100 mg iproniazid with LSD in doses of 25, 50 and 100 µg. | N = 16 (healthy) | No change to the physical or subjective effects of LSD. |
| (Fabing, 1955) | Case series | Antidepressants: Azacyclonol | 1) Pretreatment with 10-30 mg azacyclonol for 1 week before ingestion of 100 µg LSD.<br>2) Pretreatment of participants with a total of four doses of 50 mg azacyclonol at 50, 38, 26 and 2 hours before ingestion of 100 µg LSD.<br>3) Premedication with azacyclonol at either 5 or 10 mg twice daily for 7 days, followed by ingestion of 100 µg LSD. Five weeks later, participants received the same dose of LSD again without azacyclonol premedication. | 1) N = 6 (healthy) Age: 19-40<br>2) N = 2<br>3) N = 2 | 1) Azacyclonol pretreatment reduced psychological effects of LSD but has no effect on the physical effects of LSD.<br>2) The hallucinogenic effects of LSD were abolished by pretreatment of azacyclonol 50 hours before ingestion of LSD. Other neurological effects like attention deficit or physiological effects like nausea or tight sensations were still experienced by the participants<br>3) Premedication of azacyclonol for 7 days, followed by 5 weeks washout did not block the effects of LSD. One subject was administered 40 mg azacyclonol intravenously at the fifth hour post-LSD ingestion, reported decreased anxiety and decreased psychological effects. |
| (Clark, 1956) | Placebo-controlled trial | Antidepressants: Azacyclonol | 1) Pretreatment with azacyclonol for 10-40 mg for 2-5 days, followed by 100 µg LSD.<br>2) Intravenous injection of 100 mg azacyclonol was administered after 100 µg LSD. | N = 5 (schizophrenic patients) | 1) No evidence of LSD-blocking effects by azacyclonol could be observed.<br>2) No evidence of azacyclonol effects in changing the effects of LSD. |
| (Isbell & Logan, 1957) | Placebo-controlled | Antidepressants: Azacyclonol | Pretreatment with 20 mg of oral azacyclonol for 7 days and additional | N = 12 | No reduction of any aspect of LSD reaction observed from the blocking experiment. No significant diminution in any |
|  | d trial |  | 20 mg 2 hours before administration of 60 µg LSD. Additional dose of 40 mg given intravenously 3 hours after LSD administration. |  | aspect of the LSD reaction could be observed after azacyclonol administration. |
| (Barrett et al., 2000) | Cross-sectional study | Recreational: Alcohol | Self report study. Ingestion of alcohol concurrent with the administration of LSD (no more details given). | N = 22<br>Age: 18-28 years old | Ingestion of LSD concurrent with alcohol consumption diminished the effects of alcohol. No differences in subjective effects of LSD were associated with alcohol use. |
| (Preller et al., 2018) | Randomised, placebo-controlled trial | Other: Ketanserin | Pretreatment with 40 mg ketanserin 60 minutes before treatment with 100 µg LSD. | N = 24 (healthy) | Ketanserin fully blocked the subjective and mental effects of LSD, reduced communication among brain areas involved in planning and decision-making (fMRI and BOLD). |
| (Olbrich et al., 2021) | Randomised, placebo-controlled trial | Other: Ketanserin | Pretreatment with 40 mg ketanserin 60 minutes before treatment with 100 µg LSD. | N = 17 (healthy) | Ketanserin pretreatment shifted participants on LSD towards parasympathetic activity which is associated with less intense subjective experience including less audio-visual synaesthesia, ASC elementary imagery and experience of unity). |
| (Holze et al., 2021) | Randomised, placebo-controlled trial | Other: Ketanserin | Pretreatment with 40 mg ketanserin 60 minutes before treatment with 200 µg LSD. | N = 16 (healthy) | Ketanserin pretreatment followed by a high dose of LSD at 200 µg blocked subjective drug effects, ego dissolution, anxiety and oceanic boundlessness score to levels that were similar to 25 µg LSD dose. Physical effects such as blood pressure and body temperature induced by 200 µg LSD + Ketanserin were comparable to placebo controls. |
| (Becker et al., 2023) | Randomised, placebo-controlled trial | Other: Ketanserin | Oral administration of 40 mg ketanserin 60 minutes after ingestion of 100 µg LSD. | N = 24 (healthy) | Administration of ketanserin after the effects of LSD has reversed subjective and autonomic responses to LSD in humans. Duration of LSD effects were reduced from 8.5 hours to 3.5 hours (~60%) by ketanserin. Ketanserin did not alter overall mystical experiences or reversed LSD-induced increase of plasma BDNF, or alter pharmacokinetics of LSD. |
| (Resnick et al., 1965) | Placebo-controlled trial | Other: Reserpine | Pretreatment with 0.5 mg/day reserpine for 2 weeks before ingestion of 75 µg of LSD. | N = 3 (healthy) | Pretreatment with reserpine enhanced the effects of LSD on autonomic and neurologic responses as well as psychological reaction to LSD. |
| (Isbell & Logan, 1957) | Placebo-controlled trial | Other: Reserpine | Pretreatment with either 2.5 mg oral or 2 mg intramuscular reserpine 2, 10 and 22 hours prior to administration of 60 µg of LSD. | N = 12 (non-psychotic drug addicts) | Reserpine intensified LSD effects of nervousness and confusion, including nasal stuffiness, nausea, diarrhoea, vomiting and lethargy in a dose-dependent manner. Participants who underwent intramuscular administration of reserpine reported intensified LSD effects. |
| (Freedman, 1963) | Controlled trial | Other: Reserpine | Pretreatment with a single dose of 10 mg reserpine 2 days prior to 120 µg of LSD. | N = 14 (schizophrenic females) | Reserpine enhanced the physical effects of LSD including prolonged tremors and akathisia, "unpleasant" trip as well as increased duration of effects. |
| (Agnew & Hoffer, 1955) | Controlled trial | Other: Niacin | Pretreatment with 3 g of oral niacin for 3 days prior to 100 µg of LSD ingestion on test day.<br>Intravenous injection of 0.2 g niacin post-LSD ingestion (100 µg) at the height of LSD effects. | N = 10 (healthy males)<br>Age: 19-31, mean of 26 yo | Pretreatment with niacin delayed the onset of LSD-effects and prevented most of the perceptual changes from occurring.<br>Niacin administration post-LSD ingestion attenuated all effects of LSD within 5 minutes and markedly diminished the proprioceptive, perceptual, cognitive and motor effects of LSD. |
| (Isbell et al., 1959) | Placebo-controlled trial | Other: Scopolamine | Subjects were administered with either of the following four different doses of scopolamine at 0.42, 0.64, 0.85 and 1.3 mg followed by simultaneous administration of 60 µg of LSD | N = 12 | Scopolamine did not alter LSD-induced effects on patellar reflex, pupil size, blood pressure or subjective clinical effects. |
| (Isbell et al., 1959) | Placebo-controlled trial | Other: Phenoxybenzamine | Participants received different doses of phenoxybenzamine at 0.5, 1 and 1.0 mg/kg at 2 hours prior to administration of 1.0 µg/kg LSD. | N = 10 | None of the doses of phenoxybenzamine pretreatment did not alter LSD-induced effects on pulse rate, blood pressure, or subjective clinical effects. |

**Table 2.** Summary of studies and case reports describing interactions (or the absence thereof) between psilocybin and other psychoactive drugs.

| Author and year | Study type | Drug | Dose | Participants | Outcome |
| --- | --- | --- | --- | --- | --- |
| (Pokorny et al., 2016) | Randomised, placebo- controlled trial | Anxiolytics: Buspirone | Pretreatment with 20 mg oral buspirone 60 minutes before oral administration of 0.17 mg/kg psilocybin. | N = 19 (healthy) | Buspirone pretreatment markedly reduced psilocybin-induced visual perception distortion. |
| (Keeler, 1967) | Randomised, placebo-controlled trial | Antipsychotics: Chlorpromazine | Pretreatment with 50 mg chlorpromazine 2 hours before 0.2 mg/kg psilocybin. | N = 8 (healthy) | Pretreatment with chlorpromazine significantly decreased psilocybin-induced pupil dilation and visual perception distortion. |
| (Vollenweider et al., 1998) | Randomised, placebo-controlled trial | Antipsychotics: Haloperidol | Pretreatment with intravenous 0.021 mg/kg haloperidol 75 minutes before oral administration of 0.25 mg/kg psilocybin. | N = 15 (healthy) | Haloperidol pretreatment reduced psilocybin effects on the Oceanic boundlessness, measures of derealization and depersonalization phenomena but increased anxiety without any effects on visual hallucinations or delayed response task. |
| (Vollenweider et al., 1998) | Randomised, placebo-controlled trial | Antipsychotics: Risperidone | Pretreatment with either 0.5 mg or 1 mg oral risperidone 90 mins before oral administration of 0.25 mg/kg psilocybin. | N = 15 (healthy) | Risperidone pretreatment attenuated the effects of psilocybin in a dose-dependent manner. |
| (Becker et al., 2022) | Randomised, placebo-controlled trial | Antidepressants (SSRI): Escitalopram | Pretreatment with 10 mg/day for 7 days, followed by 20 mg/day for 7 days, and a single dose on test day 2 hours before 25 mg psilocybin treatment. | N = 23 (healthy) | Escitalopram pretreatment reduced negative acute effects of psilocybin (subjective bad drug effects, anxious-ego dissolution, anxiety and nadir effects). The positive mood and mind-altering effects of a full dose of psilocybin were not reduced by pretreatment. |
| (Vollenweider et al., 1998) | Randomised, placebo-controlled trial | Other: Ketanserin | Pretreatment with either 20 or 40 mg oral ketanserin 75 minutes before oral administration of 0.25 mg/kg psilocybin. | N = 27 (healthy)<br>Age: Mean 29.7 | Ketanserin pretreatment blocked psilocybin effects in a dose-dependent manner on the delayed response task and Altered State of Consciousness Rating scale. |
| (Carter et al., 2005) | Controlled trial | Other: Ketanserin | Pretreatment with 50 mg oral ketanserin 90 minutes before oral administration of 0.215 mg/kg psilocybin. | N = 8 (healthy) | Ketanserin pretreatment blocked subjective effects of psilocybin based on the 5D-ASC rating scale (anxious ego dissolution, euphoria, depersonalization and hallucination). |
|  |  |  |  |  | Ketanserin did not rescue psilocybin-induced impairment on attentional tracking performance. |
| (Carter et al., 2007) | Controlled trial | Other: Ketanserin | Pretreatment with 50 mg oral ketanserin 90 minutes before oral administration of 0.215 mg/kg psilocybin. | N = 10 (healthy) | Ketanserin pretreatment blocked the subjective effects on the 5D-ASC rating scale (anxious ego dissolution, depersonalization, euphoria and hallucination) but did not influence either the psilocybin-induced slowing of binocular rivalry or negative drug symptoms (alertness and arousal). |
| (Pokorny et al., 2016) | Randomised, placebo-controlled trial | Other: Ergotamine | Pretreatment with 3 mg oral ergotamine 100 minutes before oral administration of 0.17 mg/kg psilocybin. | N = 17 (healthy) | Ergotamine did not significantly alter subjective experiences (5D-ASC scores). |
| (Barrett et al., 2000) | Cross-sectional study | Recreational: Alcohol | Non-intoxicating levels of alcohol consumed either before or after psilocybin (self-reported, doses unknown). | N = 15 | The subjective effects of alcohol are antagonised by psilocybin, but the subjective effects of psilocybin were diminished in only 2 (out of 15) participants while only 1 reported enhanced effects, the rest (80%) reported unchanged effects. |
| (Espiard et al., 2005) | Case report | Recreational: Cannabis | Ingestion of 40 *Psilocybe semilanceata* mushrooms in infusion, followed by cannabis on the next day. | N = 1 | Psilocybin and cannabis consumption induced the hallucinogen persisting perception disorder that lasted even after discontinuation of both drugs. |

**Table 3.** Summary of studies and case reports describing interactions (or the absence thereof) between mescaline and other psychoactive drugs.

| Author and year | Study type | Drug | Dose | Participants | Outcome |
| --- | --- | --- | --- | --- | --- |
| (Lesse, 1958) | Controlled trial | Antipsychotics: Chlorpromazine and promazine | Mescaline (intravenous) or LSD (oral) followed by intravenous chlorpromazine or promazine (no more details given). | N = 25 (patients with schizophrenia) | The administration of chlorpromazine or promazine led to a reduction in acute anxiety symptoms in 68% (17/25) participants. |
| (Fabing, 1955) | Case series | Antidepressants: Azacyclonol | 1) Pretreatment with 50 mg azacyclonol at 50, 38, 26 and 2 hours before | N = 4 (healthy) | 1) Pretreatment with azacyclonol likely attenuated the effects of mescaline. |
|  |  |  | ingestion of 400 mg mescaline sulphate.<br>2) Placebo instead of pretreatment, at 4.5 or 5 hour mark 100 mg azacyclonol (intravenous). | Age: 20-45 | 2) Azacyclonol treatment during the height of the experience started to block the actions of mescaline. |
| (Clark, 1956) | Case report | Antidepressants:<br>Azacyclonol | 40 mg azacyclonol four times a day for 3 days before ingesting 200 mg and 400 mg mescaline sulphate. | N = 1<br>(healthy) | No differences in mescaline intoxication when ingesting it with or without azacyclonol pretreatment. |
| (Dittrich, 1971) | Case report | Other: 2C-O<br>(2,4,5-trimethoxy phenethylamine) | Pretreatment with 100-200 mg 2C-O and 45 min later 100 mg mescaline. | N = 1<br>(healthy)<br>Age: 29 | Pretreatment with 2C-O potentiated the effects of mescaline as the ability to concentrate decreased and time taking a test increased. |

**Table 4.** Summary of studies and case reports describing interactions (or the absence thereof) between DMT/ayahuasca and other psychoactive drugs.

| Author and year | Study type | Drug | Dose | Participants | Outcome |
| --- | --- | --- | --- | --- | --- |
| (Sai Halasz, 1963) | Controlled trial | Antidepressants (MAOI):<br>Iproniazid | Pretreated with iproniazid (4 days 100 mg/daily) followed by a pause of two days. On the following day: five volunteers received a dose of 0.65-0.83 mg/kg DMT and two individuals a lower dose of 0.35-0.55 mg/kg DMT. | N = 7<br>(healthy)<br>Age: 21-36 years<br>4 males/3 females | Volunteers who received the lower dose of DMT did not experience hallucinations or disruptions in time and space orientation, but reported an "odd" feeling. The five receiving the higher dose experienced less intense DMT effects lasting for 14-24 min. |
| (Strassman, 1996) | Randomised, placebo-controlled trial | Other: Racemic pindolol | Racemic pindolol (30 mg oral) and 0.1 mg/kg DMT (intravenous) | N = 12 (healthy) | Pindolol pre-treatment enhanced DMT effects by 2-3 times. Significant enhancement in 4-6 clinical clusters in hallucinogen rating scale. Mean arterial blood pressure effects were enhanced, heart rate responses were blunted, prolactin responses were reduced. |
| (Sai-Halasz, 1962) | Controlled trial | Other: Methysergide | Methysergide administered:<br>1) 1-2 mg perorally 30-40 min before DMT;<br>2) 0.5 mg intramuscularly 10 min before DMT. | N = 15 (healthy) | Intensification of DMT subjective effects when taken with methysergide.<br>1) Two individuals: heightened subjective effects of DMT; five individuals: very intense aggravation of DMT subjective effects.<br>2) Four individuals who took 65-80% less DMT than their first time experienced similar effects. Four individuals who took 50-60% of their first-time dose did not experience a more pronounced hallucinatory state than their first experience, and in one case, the effects were even less pronounced. |
| (Callaway & Grob, 1998) | Case report | Antidepressants (SSRI): Fluoxetine | Fluoxetine (20 mg/day) | N = 1<br>36 year old male with mild depression | Adverse effects similar to serotonin toxicity (sweating, shivering, tremors, confusion, severe nausea, vomiting and disorientation) after consumption of ~100 ml ayahuasca brew that lasted for four hours. |

**Table 5.**
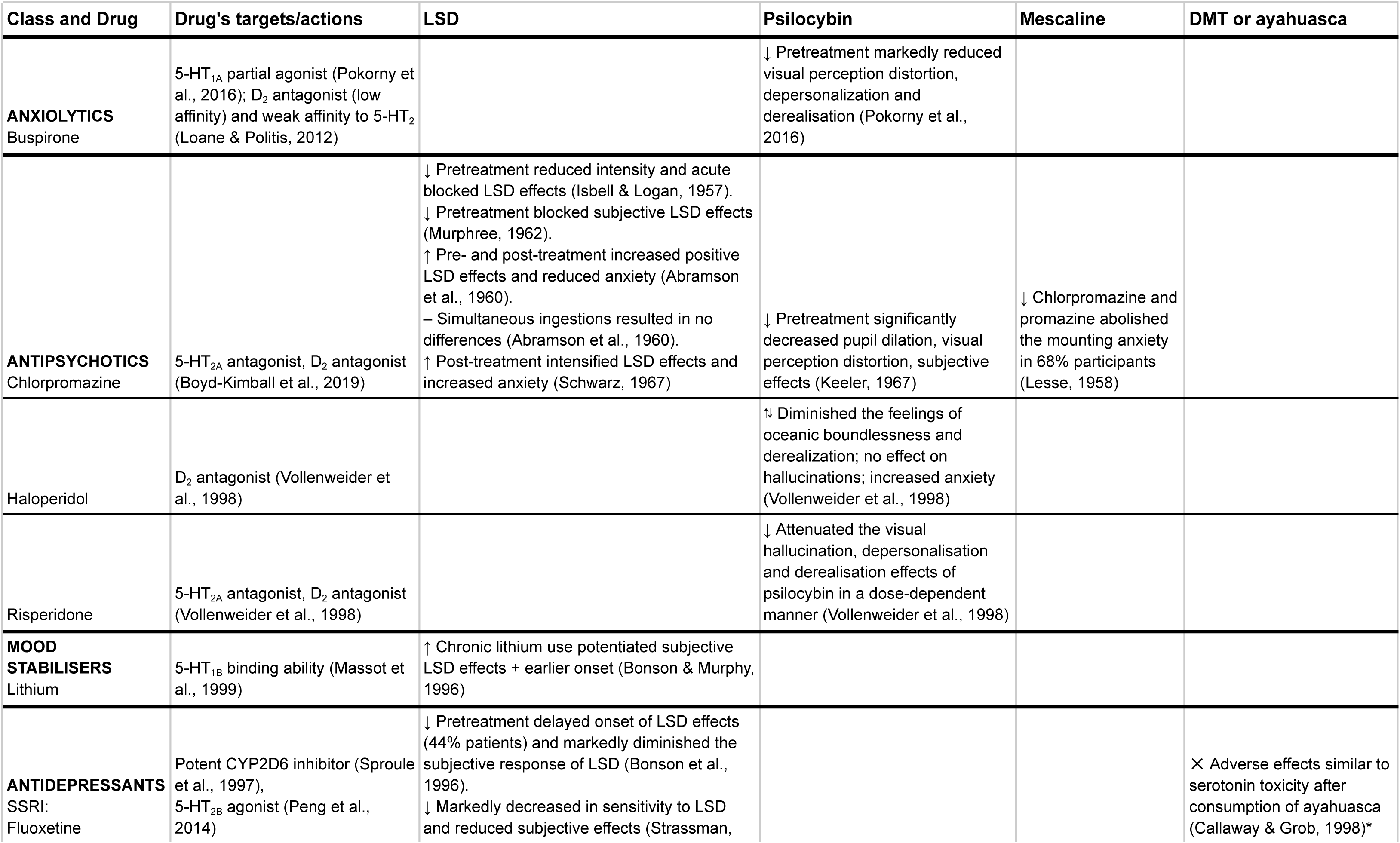
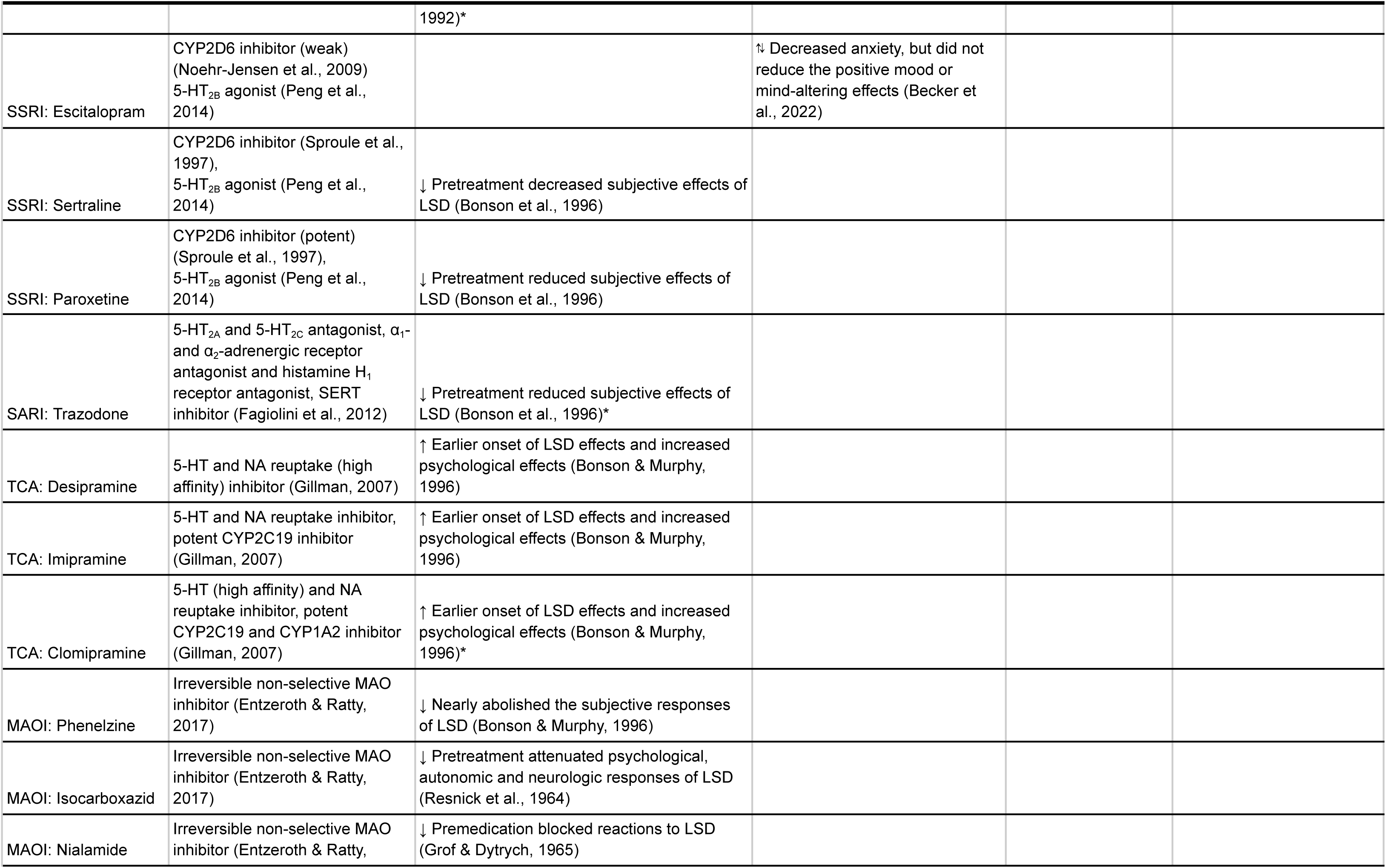
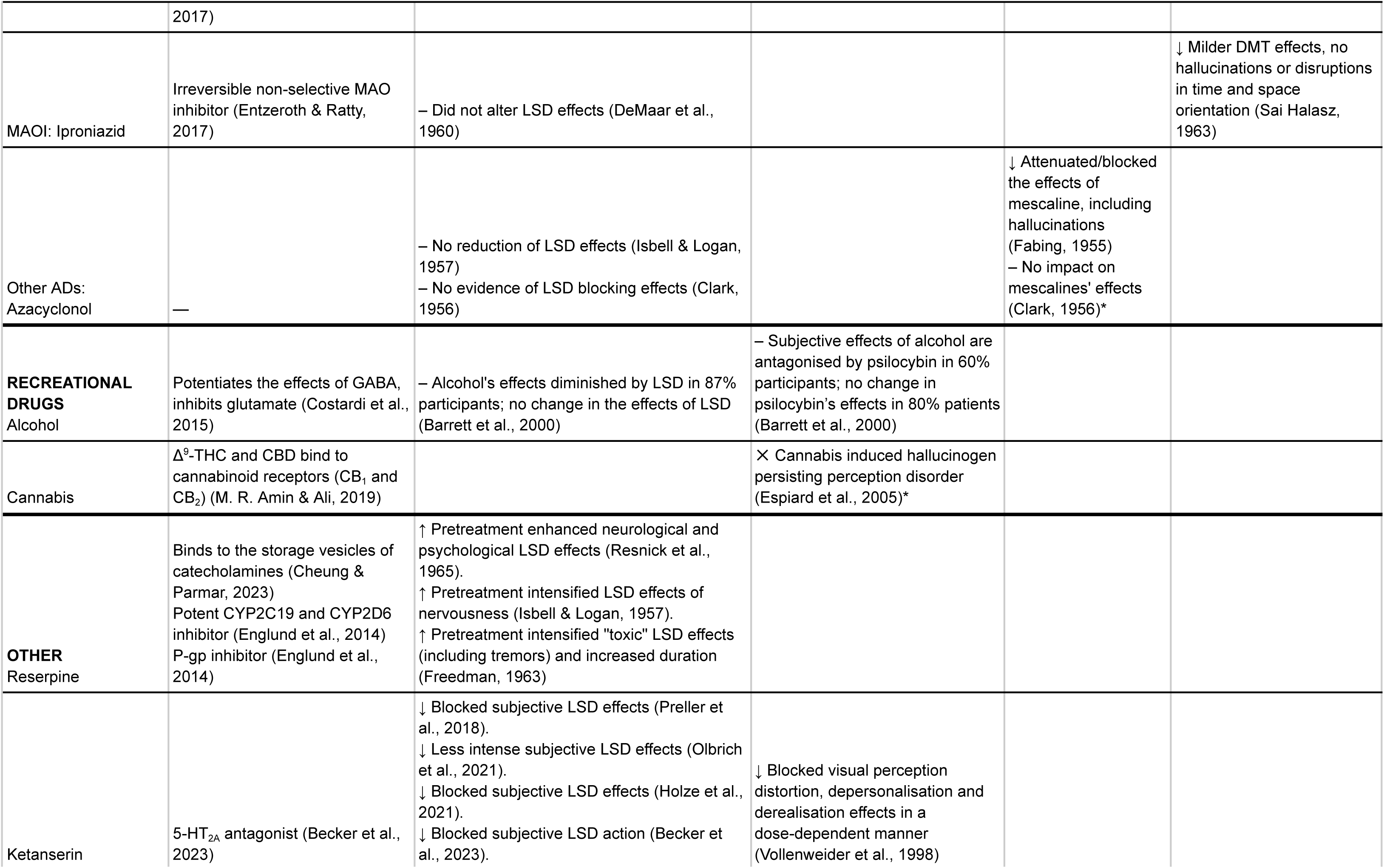
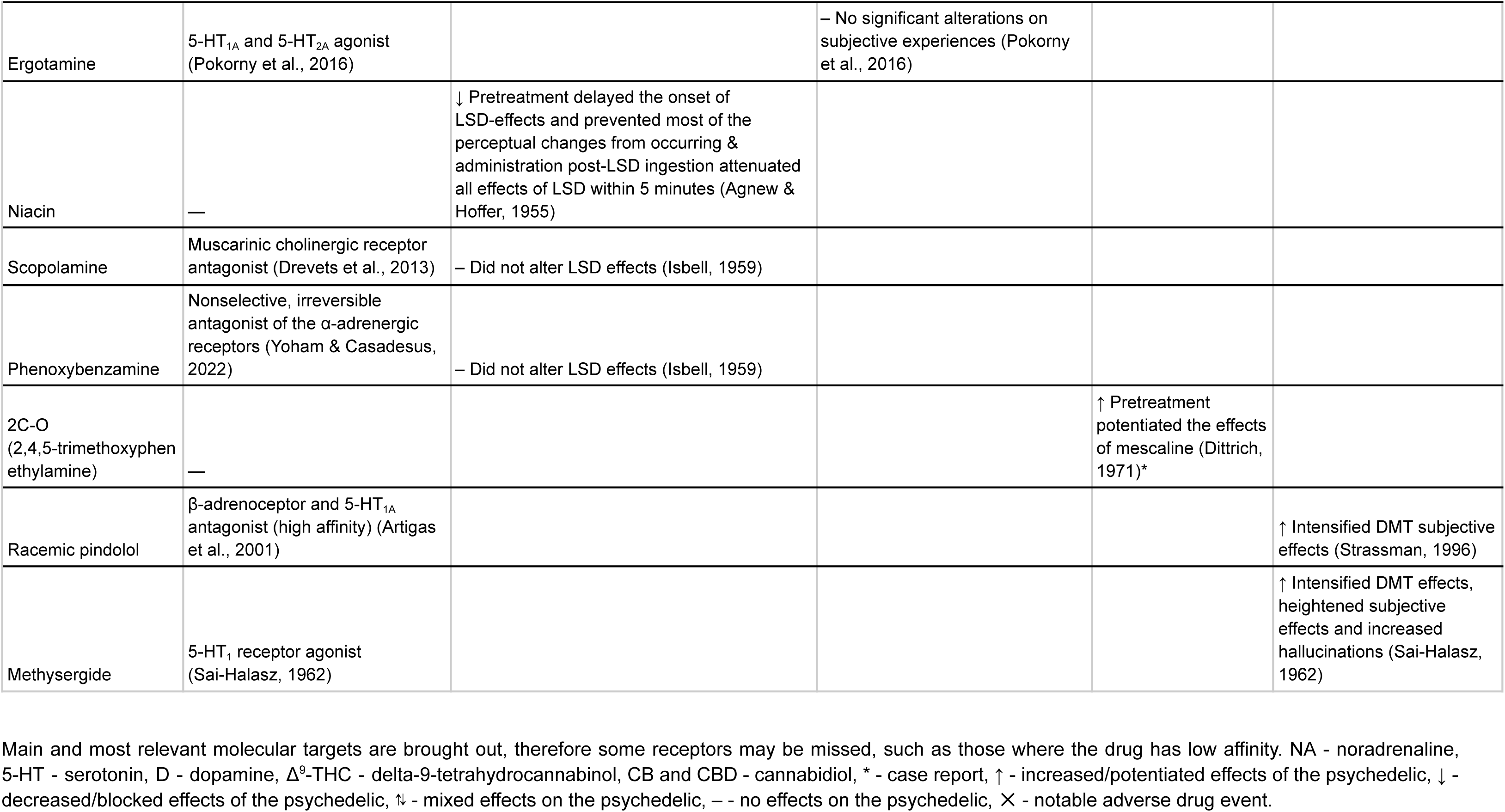
Summary table of all drug-drug interactions together with molecular targets.

## LSD

A total of 22 studies and 9 case series/reports were identified, describing interactions with antipsychotics, mood stabilisers, various antidepressants, recreational drugs and other substances. Two studies reported no effects when concomitant drugs were used.

### Antipsychotics: Chlorpromazine

A couple of studies have been done to investigate the effects of chlorpromazine pretreatment on LSD response. In a series of blocking experiments in which participants were administered 50-100 mg chlorpromazine followed by 40-60 μg of LSD ingestion, the LSD-induced subjective effects were reduced in all doses of chlorpromazine, including anxiety, changes in sensory perception and hallucination (Isbell & Logan, 1957). In the reverse experiment whereby LSD was ingested prior to oral administration of chlorpromazine, no significant difference to LSD effects were observed when compared to placebo control. However, intramuscularly-administered chlorpromazine did significantly reduce the intensity of LSD reaction (Isbell & Logan, 1957). Murphree (1962) reported contrasting results whereby pretreatment with 25 mg oral chlorpromazine just 30 minutes before was enough to block LSD-induced effects.

On the other hand, Abramson et al. (1960) reported that oral ingestion of chlorpromazine enhanced the physiological and perceptual state, as well as the hallucinations and consciousness induced by LSD, regardless of whether chlorpromazine was ingested 1.5 hours before or after LSD administration. Similarly, in a cohort of schizophrenic patients, Schwarz (1967) reported that chlorpromazine administration after LSD enhanced the effects of LSD such that one patient had thought that he had been given another dose of LSD while another subject reported feeling worse and anxious. Notably, simultaneous ingestion of chlorpromazine with LSD did not alter any of the subjective or physical effects of LSD effects (Abramson et al., 1960; Murphree, 1962).

### Mood stabilisers: Lithium

A study led by Bonson and Murphy (1996) examined the effects of lithium on LSD-effects. Three participants who had been chronically taking either lithium alone at 600 mg/day or 1000 mg/day; or in combination with the tricyclic antidepressant imipramine at 175 mg/day for 7-50 weeks stated they experienced overall enhanced effects of LSD. Additionally, there was an earlier onset of the effects along with potentiated hallucinatory and psychological effects of LSD. Each of the three participants who were interviewed reported experiencing hallucinations that were more visually vivid or intense than with the normal hallucinogenic effects of LSD which had been expected (Bonson & Murphy, 1996).

### Antidepressants (SSRI and SARI): Fluoxetine, paroxetine, sertraline and trazodone

Strassman (1992) reported a case of one adult male receiving fluoxetine to treat dysthymia. The subject had markedly decreased sensitivity to LSD whereby the usual dose that was capable of eliciting a full hallucinogenic effect was only ½ or ⅓ as effective in producing the desired effects after pretreatment with fluoxetine (Strassman, 1992). In 1996, a cohort study was conducted by Bonson et al. (1996) to investigate the effects of various SSRIs including fluoxetine, sertraline, paroxetine and serotonin receptor antagonists and reuptake inhibitors (SARI) trazodone, on the LSD response. In general, pretreatment of at least 2 weeks with SSRIs or SARI attenuated the overall response to LSD such as decreased physical, hallucinogenic and psychological effects of LSD regardless of dose or duration of pretreatment with the SSRI. Notably, one subject who had been on fluoxetine for 1 week reported attenuated effects of LSD and after a month of fluoxetine discontinuation, the normal responses to LSD returned (Bonson et al., 1996).

### Antidepressants (TCA): Imipramine, desipramine and clomipramine

The effects of multiple tricyclic antidepressants including imipramine, desipramine and clomipramine on LSD response were investigated by Bonson and Murphy (1996). Participants were treated with 125 mg to 200 mg/day tricyclic antidepressants for at least 8 weeks prior to administration of LSD at a minimum dose of 80 μg. The pretreatment of the TCAS led to an earlier onset of LSD effects as well as increased the physical, hallucinatory and psychological effects of LSD. Notably, one patient who had stopped desipramine treatment for 12 weeks still experienced the potentiated effects of LSD in terms of psychological and hallucinatory response but not at 20 weeks after. Conversely, another patient who had stopped clomipramine treatment for 12 weeks no longer felt the enhanced effects of LSD (Bonson & Murphy, 1996).

### Antidepressants (MAOI): Isocarboxazid, phenelzine, nialamide and iproniazid

Three independent studies have investigated the effects of MAOI on LSD response. Resnick et al. (1964) were amongst the first to investigate the effects of MAOI isocarboxazid (brand name Marplan). Pretreatment of participants with 30 mg/day isocarboxazid for 5 days or 2 weeks prior to oral administration of 40 μg LSD attenuated autonomic and psychological responses of LSD (Resnick et al., 1964). Similarly, Bonson and Murphy (1996) reported that pretreatment of participants with 60-75 mg/day phenelzine for 12 weeks followed by 150 μg of LSD dose nearly abolished subjective responses to LSD (Bonson & Murphy, 1996). Pretreatment with nialamide, a different MAOI, daily for 3 weeks to reach a total dose of 3500 mg also blocked the effects of LSD and patients tolerated very high doses of LSD at 500 μg without any clinical or psychotic symptoms (Grof & Dytrych, 1965). In contrast, iproniazid at 100 mg, when taken together with varying doses of LSD, did not alter any physical or subjective effects of LSD (DeMaar et al., 1960).

### Recreational drugs: Alcohol

There are very limited studies on the effects of alcohol on LSD response. A retrospective self-report study conducted by Barrett et al. (2000) aimed to explore the effects of alcohol and LSD on each other when consumed together. A total of 13 out of 15 participants (86.7%) reported complete blockage of subjective alcohol effects while under the influence of LSD and the remainder reported diminished effect of alcohol. These participants reported that higher doses of alcohol were required to become intoxicated when under the influence of LSD. One of them also noticed that none of the alcohol effects could be felt until the effects of LSD had worn off and if LSD was taken after feeling intoxicated, the alcohol effects would dissipate after LSD intake. In regards to the effects of alcohol on LSD however, none of the participants observed any consequence by alcohol on the subjective effects of LSD (Barrett et al., 2000).

### Other: Reserpine

Reserpine was found to enhance some aspects of LSD-effects in a few studies (Freedman, 1963; Isbell & Logan, 1957; Resnick et al., 1965), with a dose-dependent effect observed in one study. Isbell and Logan (1957) first investigated the effects of reserpine in twelve non-psychotic adults in a series of experiments with varying concentrations of reserpine and administration route. Participants pretreated with either a single dose of 1 mg oral reserpine 2 hours prior to 60 μg LSD; 5 mg oral reserpine divided over 2 doses at 10 and 2 hours prior to 60 μg LSD treatment did not experience any blocking effects from reserpine. However, participants treated with 7.5 mg oral reserpine divided over three doses at 22, 10 and 2 hours prior to LSD treatment experienced worsened effects (Isbell & Logan, 1957).

Leaving aside the expected reserpine side-effects such as nasal stuffiness, nausea, diarrhoea, vomiting, lethargy, weakness and dizziness, individuals also had increased nervousness and confusion when used LSD together with reserpine. Over half of the patients who received 6 mg of reserpine intramuscular with 60 μg/kg of LSD had gross tremors at rest. Such tremors did not occur when the same dose of LSD was taken alone in these patients (Isbell & Logan, 1957).

When participants were pretreated with a lower dose of 6 mg reserpine in total divided over three doses in the same interval via intramuscular injection prior to 60 μg LSD, they experienced increased irritability and mood depression as well as enhanced neurological changes (Isbell & Logan, 1957). Interestingly, the combination of LSD and reserpine was reported to cause a specific type of hallucination called “jets” or “jet propulsion.” This hallucination is characterised by a hissing sound starting from the back of the head and culminating in a flash of light, along with a sensation of flying or being hurled through the air. This hallucination has not been observed when LSD or reserpine is taken alone. Notably, participants that had the same dose of reserpine via intramuscular injection against a lower dose of LSD at 0.5 μg/kg experienced more positive subjective effects, as reflected in the increased number of LSD-related positive answers and clinical grade, without gross tremors (Isbell & Logan, 1957).

Freedman (1963) conducted a study of 14 adult schizophrenic women where the participants were given a single dose of 10 mg reserpine, 2 days prior to 120 μg LSD on test day. All of the participants described that the effects lasted longer and were overall more unpleasant than the control situation without reserpine pretreatment. The experimenters also observed that most of the participants experienced extended periods of tremor and akathisia and in one case, suffered an oculogyric crisis (Freedman, 1963).

Finally, Resnick et al. (1965) examined the effects in three healthy adult males who were treated with 0.5 mg/day of reserpine for two weeks before ingesting 75 μg of LSD. They found that pretreatment with reserpine markedly enhanced the effects of LSD on autonomic and neurologic responses as well as psychological reaction to LSD. Autonomic responses such as blood pressure, heart rate and pupillary size were enhanced and neurologic functions such as cerebellar functions and deep tendon and periosteal reflexes were affected. The psychological reaction to LSD was also enhanced, as indicated by interview, behavioural observation and a symptom rating scale (Resnick et al., 1965).

### Other: Azacyclonol and scopolamine

Fabing (1955) conducted a number of experiments to investigate the effect of azacyclonol (brand name Frenquel) on LSD-induced responses. Participants were pretreated with 10-30 mg/day of oral azacyclonol for 1 week and observed that the hallucinatory, conscious and mood-altering effects of LSD were blocked but some small physiological effects such as muscle twitching and nausea remained. They followed up and reported that similar blocking effects were also observed when the participants were pretreated with four doses of 50 mg azacyclonol from 50 hours before the test (Fabing, 1955). They then sought to determine whether the blocking effects of azacyclonol persist after treatment had stopped. The two participants in that experiment reported normal LSD-experiences. One of the participants was administered 40 mg azacyclonol intravenously at the fifth hour post-LSD ingestion and felt less anxious while he realised that he was ‘back in the real world’ again and was able to follow conversations more easily (Fabing, 1955). An independent study by Isbell and Logan (1957) reported no changes in LSD-induced effects were observed in participants treated with 20 mg of azacyclonol 3 times a day for 1 week. A reversal experiment whereby intravenous administration of azacyclonol of 100 mg in divided doses after LSD did not alter the intensity of LSD-reactions (Isbell & Logan, 1957). Similarly, Clark (1956) did not observe any blocking of LSD-effects when the participants were pretreated with Frequel varying from 10 to 40 mg, four times daily for 2 to 5 days. Azacyclonol, when administered intravenously at the height of LSD reaction, did not alter the experience or the duration of effects (Clark, 1956).

The effects of another antidepressant scopolamine were also investigated whereby various doses of scopolamine were subcutaneously administered simultaneously with 1 μg/kg LSD (Isbell et al., 1959). The side effects of scopolamine including pupil dilation, dry mouth and blurred vision increased in a dose-dependent manner and some were more prominent in combination with LSD than LSD alone. Assessment of the mental effects of scopolamine was more complicated. Even though there were an increased number of positive answers after the combination of LSD and scopolamine when compared to LSD alone, the specific responses linked to the positive answers, such as sleepiness and vision blurring, were attributed to scopolamine use alone. The frequency of LSD-specific reactions such as visual perception distortion and optical hallucination were no different in LSD and scopolamine combination when compared to LSD alone (Isbell et al., 1959).

### Other: Ketanserin

Few recent studies have investigated the effects of serotonin receptor antagonist ketanserin on the autonomic nervous system following LSD administration. Preller et al. (2018) conducted a study in 24 healthy participants, whereby participants were pretreated with 40 mg oral ketanserin at three different doses two weeks apart with the final dose 60 minutes before treatment with LSD. Neuroimaging scans performed at 75 and 300 minutes after treatment administration showed that ketanserin blocked the global brain connectivity induced by LSD. Notably, the global brain connectivity in somatomotor network is correlated with subjective effects of LSD (Preller et al., 2018). In a double-blind placebo controlled trial, ketanserin pretreatment followed by LSD administration led to a significant shift towards the parasympathetic activity when compared to placebo only or placebo-LSD controls (Olbrich et al., 2021). The shift in autonomic nervous system was also associated with less intense subjective experience of LSD measured by the Five Dimensional Altered States of Consciousness (5D-ASC) scale (Olbrich et al., 2021). Another independent study by Holze et al. (2021) reported that ketanserin pretreatment blocked the subjective effects of a high dose of 200 μg LSD to levels similar to 25 μg of LSD. The scores for “Anxious Ego-Dissolution” and “Oceanic Boundlessness” as well as any subjective “good” and “bad” drug effects at 200 μg LSD + Ketanserin were similar to 25 μg of LSD. They also observed that the combination significantly lowered the blood pressure, body temperature and heart rate of the participants to similar levels as in placebo controls (Holze et al., 2021). The blocking effects of ketanserin on LSD were also corroborated by Becker et al. (2023) who observed that the participants pretreated with 40 mg ketanserin followed by 100 μg of LSD one hour later, experienced less LSD-induced auditory and visual alterations as well as ego-dissolution and emotional excitement. However, ketanserin pretreatment did not significantly alter overall mystical experiences or plasma brain-derived neurotrophic factor (BDNF) levels induced by LSD. Ketanserin-pretreated participants also experienced shortened duration of LSD-effects from 8.5 to 3.5 hours without any change in pharmacokinetics of LSD (Becker et al., 2023).

### Other: Niacin

Agnew and Hoffer (1955) examined the effects of niacin (vitamin B3) on the effects of LSD. Ten healthy adults were administered 200 mg niacin intravenously post LSD administration. Attenuation of LSD effects were observed in all participants and niacin injection markedly diminished the proprioceptive, perceptual and cognitive effects of LSD. However, when participants were pretreated with 3 grams of oral niacin daily for 3 days followed by 100 μg of LSD on test day, the onset of LSD effects was delayed and participants still experienced feelings of unreality and depersonalisation, but without LSD-induced perceptual changes (Agnew & Hoffer, 1955).

### Other: Phenoxybenzamine

Isbell et al. (1959) examined the blocking effects of varying doses of phenoxybenzamine pretreatment at 0.5 mg, 1 mg and 1.0 mg/kg that were administered 2 hours prior to 1 μg/kg LSD. None of the doses of phenoxybenzamine significantly altered LSD-induced effects including pupil dilation, pulse rate, blood pressure, knee reflex or subjective positive feelings (Isbell et al., 1959).

### Additional case reports

Additionally, there is a report about a 16 year old individual who consumed two doses of LSD while being on fluoxetine (20 mg/day) treatment and developed marked stupor hours after LSD ingestion, followed by focal seizure that progressed to a grand mal convulsion (Picker et al., 1992). However, this report is omitted from our results as the person previously ingested LSD 30 times during fluoxetine treatment without such adverse effects and it is unclear whether the person had any previous experiences with the same batch of LSD.

## Psilocybin

A total of 10 studies and 1 case report were identified, describing interactions with anxiolytics, antipsychotics, antidepressants, recreational drugs and other substances.

### Anxiolytics: Buspirone

A study conducted by Pokorny et al. (2016) pretreated 19 healthy participants with 20 mg buspirone one hour before the oral ingestion of 0.17 mg/kg psilocybin. Buspirone pretreatment markedly reduced the psilocybin-induced visual hallucinations and to a lesser extent, facilitated imagination but did not affect audio-visual synaesthesia. Additionally, it attenuated the psilocybin-induced loosening of ego-boundaries, changes in sense of time and euphoria (Pokorny et al., 2016).

### Antipsychotics: Chlorpromazine

In one study by Keeler (1967), eight healthy volunteers were given either 50 mg of chlorpromazine or placebo two hours before oral administration of 0.2 mg/kg psilocybin. They reported that the participants pretreatment with chlorpromazine experienced significantly decreased psilocybin-induced pupil dilation, visual perception distortion and after-image response (Keeler, 1967).

### Antipsychotics: Risperidone and haloperidol

In a randomised, placebo-controlled trial conducted by Vollenweider et al. (1998), it was reported that both risperidone and haloperidol reduced the effects of psilocybin. Risperidone affected psilocybin in a dose-dependent manner, with 0.5 mg blocking the effects of psilocybin by 69-78% and 1 mg completely abolishing psilocybin-induced psychosis (98-99%). Haloperidol pretreatment diminished the feelings of oceanic boundlessness and derealization, without affecting visual perception distortions or hallucinations. However, participants who underwent haloperidol treatment experienced increased anxiety and dread of ego-dissolution (Vollenweider et al., 1998).

### Antidepressant: Escitalopram

Becker et al. (2022) conducted a randomised placebo-controlled trial involving 23 healthy participants with two experimental test sessions to examine the interaction effects between the antidepressant escitalopram and psilocybin. Briefly, participants were pretreated with 10 mg escitalopram daily for 7 days, followed by 20 mg/day for another 7 days including test day which was ingested 2 hours before 25 mg psilocybin administration. They observed that pretreatment with escitalopram significantly reduced psilocybin-induced ego disintegration and anxiety, but had no effect on depersonalisation, oceanic boundlessness, or euphoria. There were also no significant differences between the groups in overall mystical experiences induced by psilocybin although it was able to reduce acute adverse effects on blood pressure. Notably, escitalopram did not normalise psilocybin-induced increase in plasma BDNF (Becker et al., 2022).

### Other: Ketanserin

In a separate experiment, Vollenweider et al. (1998) pretreated 25 healthy volunteers with 20 mg of oral ketanserin or 40 mg of placebo, followed by 0.25 mg/kg psilocybin 75 minutes later. It was reported that ketanserin blocked psilocybin-induced psychosis in a dose-dependent manner. Pretreatment with 20 mg of ketanserin reduced the altered state of consciousness induced by psilocybin by 50-70%, while 40 mg of ketanserin completely prevented the effects of psilocybin in 4 out of 5 participants. In the remaining participant, the effects of psilocybin were markedly reduced by 75-87% (Vollenweider et al., 1998). Similarly, Carter et al. (2005) pretreated 8 healthy participants with 50 mg oral ketanserin 90 minutes prior to 0.215 mg/kg psilocybin and examined the effects on attentional tracking ability and spatial working memory as well as subjective effects. Ketanserin significantly blocked the subjective effects of psilocybin including “Oceanic Boundlessness” (depersonalisation, euphoria), “Anxious Ego Dissolution” (ego-disintegration, anxiety) and “Visual Restructuralization” (hallucinations, synaesthesia) but not “Auditory alterations” or “Reduction of Vigilance” which measures acoustic hallucinations and reduced alertness respectively. However, ketanserin pretreatment did not rescue the psilocybin-induced decrease on attentional tracking ability, whereby participants were to track specific coloured disk on the monitor along with other coloured disks, the disks were then all coloured the same during the prompt and the user has to select the correct disk of a target colour (Carter et al., 2005). They followed up with another experiment (Carter et al., 2007) with the same experimental design but examined the effects on binocular rivalry whereby participants were presented with two incongruent images which typically results in perceptual alternation between the two images. Pretreatment with ketanserin did not alter the psilocybin-induced slowing of binocular rivalry. However, ketanserin pretreatment has consistently lowered the psilocybin-induced effects on Oceanic Boundlessness, Anxious Ego Dissolution and Visual Restructuralization scores without affecting the “Reduction of Vigilance” score as well as the arousal score which is a new addition to a more recent version of 5D-ASC (Carter et al., 2007).

### Other: Ergotamine

Pokorny et al. (2016) conducted a study whereby 17 participants were pretreated with 3 mg ergotamine 100 minutes before the oral administration of 0.17 mg/kg. They observed that ergotamine pretreatment did not significantly alter any of the psilocybin-induced effects including sense of euphoria, alteration to sense of time, anxious ego disintegration, or visual hallucinations based on the 5D-ASC score (Pokorny et al., 2016).

### Recreational drugs: Alcohol and cannabis

Barrett et al. (2000) conducted a retrospective self-report study to examine the effects of non-intoxicating levels of alcohol on psilocybin when consumed either before or after psychedelic ingestion. Fifteen participants underwent structured interviews consisting of a series of standardised open-ended questions: “Have you ever used alcohol while under the influence of psychedelic or psychedelic while under the influence of alcohol?”, “Did you notice any difference in the effect of either the alcohol or psychedelic when you used the two together?”,“What was the nature of the difference?”. The answers were rated on a five-point Likert scale by the investigators. The subjective effects of alcohol are partially antagonised by psilocybin whereby users reported diminished response to alcohol but none of the users reported complete blocking of alcohol effects. However, 80% of the participants did not experience any difference in effects of psilocybin when consumed together with alcohol (Barrett et al., 2000).

Espiard et al. (2005) reported a case of an 18-year old student who was hospitalised for chronic perceptual impairments and dysphoric mood lasting for 8 months. Further questioning revealed that the perceptual distortions initially appeared after ingestion of infusion made of 40 *Psilocybe semilanceata* mushrooms, which appeared again the next day during a cannabis consumption. The symptoms faded after the discontinuation of cannabis use but then increased again 6 months later. The subject was then diagnosed with hallucinogen persisting perception disorder (HPPD). The authors concluded that psilocybin consumption led to the hallucinatory experience in the chronic cannabis smoker and continuation of cannabis consumption could potentially stimulate the reappearance of the symptoms (Espiard et al., 2005).

## Mescaline

A total of 1 study and 3 case series/reports were identified, describing interactions with antipsychotics, antidepressants and other compounds.

### Antipsychotics: Chlorpromazine

In one study, 25 schizophrenic patients received either mescaline by intravenous injection or oral LSD, followed by chlorpromazine or promazine intravenous injection (Lesse, 1958). The authors noted that the anxiety caused by mescaline and LSD administration was abolished in 68% (17 out of 25) of the participants by chlorpromazine or promazine. No details were given on the number of patients who received mescaline or LSD or on the doses and timing of administration.

### Antidepressants: Azacyclonol

Another study investigated azacyclonol (Frenquel) as a blocking agent for mescaline effects (Fabing, 1955). They used four healthy patients, two of whom received pretreatment with 50 mg azacyclonol at 50, 38, 26 and 2 hours before ingesting 400 mg of mescaline sulphate, while the other two received placebo pretreatment and 100 mg intravenous injections of azacyclonol at 4.5 and 5 hours, respectively. The patients who received azacyclonol as pretreatment developed nausea and minor subjective effects (such as feeling tension in leg muscles and disruptions in thought continuity), but did not experience hallucinations or other psychic accompaniments. One of the other two patients experienced euphoria and sensations of levitation with brightened colours, while the other two developed somatic hallucinations and extreme coldness (vomited at the 4^th^ hour). After receiving a 100 mg intravenous treatment of azacyclonol, the effects of mescaline were blocked, and both patients returned to “normal” within an hour.

### Case reports

A self-experiment was conducted by Clark (1956), who took azacyclonol four times a day for three days before ingesting 200 mg and 400 mg of mescaline sulphate. He reported that the intoxications were identical in all respects to those he previously experienced after taking the same doses without azacyclonol pretreatment, and did not experience any blocking effect of azacyclonol (Clark, 1956).

Another self-experiment conducted by Dittrich (1971), who used mescaline in combination with 2,4,5-trimethoxyphenethylamine (2C-O). He designed a blind experiment in which he first ingested 300 mg of 2C-O, then 100 mg and 200 mg of 2C-O with mescaline, and finally 250 mg of mescaline alone. He did not experience any subjective effects while taking 2C-O alone. In regards to the experiment with pretreatment, he observed differences in two tests. Mescaline was shown to decrease performance in a test measuring concentration on optical details, while pretreatment with 2C-O resulted in even greater deterioration of performance than mescaline alone. The time to complete a test (semantic differential) was measured and increased under mescaline compared to placebo and even further after pretreatment with 2C-O (Dittrich, 1971).

## DMT and ayahuasca

A total of 3 studies were identified for DMT, describing interactions with antidepressants as well as other substances. One case report was found for ayahuasca.

### Antidepressants (MAOI)

In an experimental study by Sai Halasz (1963), seven volunteers (four males and three females aged between 21-36 years) were pre-treated with the MAOI iproniazid for four days, followed by a two-day washout period to eliminate the direct effect of iproniazid while preserving its MAOI activity. On the fifth day, two individuals received a reduced dose of DMT (0.35-0.55 mg/kg), while the other five received a dose of 0.65-0.83 mg/kg.

The individuals who received the reduced dose of DMT reported no significant changes in their perception of time, space, or sensory experiences. However, they reported an “odd” or “strange” feeling, which they found difficult to describe. The five volunteers who received the higher DMT dose experienced two distinct phases of effects. The first phase lasted 14-24 minutes and was similar to the effects of DMT, albeit less pronounced. They reported seeing a few hallucinations, which lacked vivid colours and only occurred with their eyes closed. In the second phase, the hallucinations disappeared and their perception of time and space returned to baseline levels. However, the feeling of oddness remained and even intensified (Sai Halasz, 1963).

### Other: racemic pindolol and methysergide

In a four-cell double-blind randomised study by Strassman (1996), twelve volunteers received a sub-hallucinogenic dose (0.1 mg/kg) of intravenous DMT or saline placebo, in combination with either 30 mg of oral racemic pindolol or placebo-pindolol. The results of the study showed that pindolol (a potent 5-HT_1A_ antagonist) pre-treatment enhanced DMT effects by two to three times, which was substantiated by scores on the hallucinogen rating scale. Four to six clinical clusters (affect, volition, somatic effects, perception, cognition and intensity) demonstrated a significant enhancement by pindolol. Heart rate responses were blunted, probably due to pindolol’s anti-sympathetic effects, while mean arterial blood pressure effects were enhanced. Prolactin responses were reduced, while those of adrenocorticotropin were unaffected (Strassman, 1996).

In a quasi-experimental study by Sai-Halasz (1962), 40 patients (20 males and 20 females aged between 18-44 years) received a DMT dose of 0.76-1.03 mg/kg. Two to three months later, 15 of those individuals participated in the second experiment. Seven of them were administered the same dose as the first time (0.81-0.89 mg/kg DMT), while eight patients had their dose reduced to 50-80% of their first dose. In the second experiment, the participants were pre-treated with methysergide (1-methyllysergic acid butanolamide), a compound structurally similar to LSD and a serotonergic antagonist but without psychic effects. Methysergide was administered perorally (1-2 mg) 30-40 min before DMT or intramuscularly (0.5 mg) 10 min before DMT. Out of the seven patients who received the same DMT dose as the first time, five individuals reported very intense aggravation of DMT subjective effects, such as more intense hallucinations, more brilliant colours, deeper loss of time and perception and lost all relation with the world, unlike the first time. The other two individuals reported heightened subjective effects of DMT, but not to such a marked degree. Four individuals who took 65-80% less DMT than their first time experienced similar effects as their first experience. The four individuals who took 50-60% of their first-time dose did not experience a more pronounced hallucinatory state than their first experience. In one case, the effects were even less pronounced (Sai-Halasz, 1962).

### Case reports

There was one case report by Callaway and Grob (1998) of a 36-year-old male individual who was receiving fluoxetine treatment (20 mg/day) and who participated in a ceremonial ayahuasca session. Approximately one hour after ingesting about 100 ml of ayahuasca, the patient developed sweating, shivering, tremors and confusion, which gradually progressed and led to severe nausea, vomiting and disorientation. The patient did not receive any treatment and the symptoms subsided after four hours following the ingestion of ayahuasca (Callaway & Grob, 1998).

Additionally, another case report by Bakim et al. (2012) involved a 42 year old male who received a combination of quetiapine at a daily dose of 1000 mg and fluoxetine at a daily dose of 40 mg. While the individual did not consume ayahuasca and therefore not included in the main results, he did ingest a spoonful of harmal (*Peganum harmala*) to treat his haemorrhoids, a plant used in ayahuasca brew. Two hours after ingestion of harmal the individual developed symptoms such as nausea, vomiting, sweating, tremors, followed by confusion with visual hallucinations, agitation and loss of orientation and was diagnosed with serotonin syndrome. Symptoms resolved within 24 hours after starting the treatment with cyproheptadine, chlorpromazine and diazepam (Bakim et al., 2012).

## Discussion

In this systematic review, a total of 8,487 records were screened and 34 reports from 50 studies were included, providing information about the potential drug-drug interactions (or the lack of) of classic psychedelics in the scientific literature. Considering the scarcity of studies addressing DDIs involving classic psychedelics, case reports were included to provide more insights into DDIs. Although anecdotal in nature, these case reports highlight potential DDIs and areas for future research.

Classic psychedelics are known to bind to 5-HT_2A_ receptors where their hallucinogenic properties are thought to be mediated with (Vollenweider & Smallridge, 2022). The results of multiple studies on drugs that are 5-HT_2A_ receptor antagonists, such as ketanserin, trazodone, risperidone and chlorpromazine, indicated that they were all effective in attenuating the effects of LSD and psilocybin. Specifically, **ketanserin**, a 5-HT_2A_ antagonist with a strong affinity (Becker et al., 2023) was found in all studies to either fully block or reduce subjective effects of LSD (Becker et al., 2023; Holze et al., 2021; Olbrich et al., 2021; Preller et al., 2018) and psilocybin (Carter et al., 2005, 2007; Vollenweider et al., 1998). Similarly, decreased physical effects were also observed, including lowered blood pressure, body temperature and heart rate (Holze et al., 2021). Interestingly, while it has been shown in one study that 5-HT_2A_ blocking with ketanserin diminished the hallucinogenic, depersonalization and derealization effects of LSD, it did not affect the mystical experiences, suggesting that these effects could be modulated by other mechanisms (Becker et al., 2023). Similarly, **risperidone**, a 5-HT_2A_ but also D_2_ antagonist was found to attenuate effects of psilocybin in a dose-dependent manner (Vollenweider et al., 1998). **Trazodone**, another 5-HT_2A_ antagonist (Balsara et al., 2005), was shown to reduce the psychological and hallucinogenic effects of LSD in one case report (Bonson et al., 1996).

The findings from studies using **chlorpromazine**, which is also a 5-HT_2A_ receptor antagonist but has affinity for D_2_ and adrenoceptor α_1A_ and α_1B_ subtypes as well (Boyd-Kimball et al., 2019; Gillman, 1999), are inconsistent. While two studies (Isbell & Logan, 1957; Murphree, 1962) found that chlorpromazine reduced the intensity of LSD-induced effects as well as anxiety, another two independent studies (Abramson et al., 1960; Schwarz, 1967) reported the opposite. In addition, while Isbell and Logan (1957) reported that oral pre-administration of chlorpromazine blocked the effects of LSD and not post-administration, then Abramson and colleagues (1960) observed enhanced effects regardless of whether it was administered before or after LSD intake. However, the inconsistent findings regarding the effects of chlorpromazine on LSD response may be due to differences in study design, dosing regimens and individual variability. Moreover, chlorpromazine was reported to have blocking action also for psilocybin and mescaline, where its pretreatment significantly decreased psilocybin-induced visual perception distortion (Keeler, 1967) and abolished the increased anxiety observed in schizophrenic patients following mescaline ingestion (Lesse, 1958).

Pretreatment with **buspirone**, which is a 5-HT_1A_ partial agonist, markedly reduced the psilocybin-induced visual hallucinations (Pokorny et al., 2016). On the contrary, agonism of 5-HT_1A_ with **ergotamine** did not affect any of the psilocybin effects (Pokorny et al., 2016). However, given that ergotamine has lower efficacy at receptor signal transduction when compared to buspirone and its general low bioavailability, it was suggested that the administered dose may not have been high enough to compete with psilocybin at those receptor site (Pokorny et al., 2016). These findings indicate that 5-HT_1A_ receptors may be also involved in the manifestation of psilocybin-induced effects such as visual hallucinations, affective alterations, derealization and depersonalization. The mechanism of action by which buspirone reduces hallucinations may involve direct stimulation of 5-HT_1A_ receptors, or alternatively, interaction between 5-HT_1A_ and 5-HT_2A_ receptors on pyramidal cells due to co-expression of 5-HT_1A_ and 5-HT_2A_ receptors in cortical and visual areas (Pokorny et al., 2016). Despite both psilocin and buspirone being partial agonists at 5-HT_1A_ receptors, the blocking effect of buspirone may be due to a more effective inhibitory impact on pyramidal neurons compared to psilocin (Pokorny et al., 2016).

Buspirone is also an antagonist for dopamine D_2_ receptors (low affinity) and has shown weak affinity to the 5-HT_2_ receptors (Loane & Politis, 2012), which could have a role in modulating the effects of psilocybin as well. However, while LSD is known to have an effect on the dopaminergic system (found to be important on perceived selfhood and cognition) (Lawn et al., 2022) and it has high affinity for both D_1_ and D_2_ receptors (Halberstadt & Geyer, 2011), then psilocin does not bind to dopamine receptors (Halberstadt & Geyer, 2011). Moreover, D_2_ antagonism by haloperidol alone did not have an effect on psilocybin-induced hallucinations (Vollenweider et al., 1998). **Haloperidol** did, however, diminish the feelings of oceanic boundlessness and derealization and also increased anxiety (Vollenweider et al., 1998). In this case, psilocybin may exert an indirect impact on dopaminergic systems, which is subsequently counteracted by haloperidol (Vollenweider et al., 1998). It has been shown in rats that psilocybin administration can increase extracellular dopamine levels in the frontal cortex (Wojtas et al., 2022) and psilocin administration has also increased dopamine levels in the nucleus accumbens (Sakashita et al., 2015).

Interestingly, blocking of serotonin reuptake transporters with SSRIs, such as **fluoxetine**, **sertraline** and **paroxetine,** reduced the effects of LSD as reported in one study (Bonson et al., 1996). Moreover, fluoxetine also delayed the onset of LSD effects in nearly half of the participants in the same study (Bonson et al., 1996) and was shown to markedly decrease sensitivity to LSD in one individual (Strassman, 1992). Furthermore, trazodone, which was previously described to reduce LSD effects as it is a 5-HT_2A_ antagonist, is also a serotonin receptor antagonist and reuptake inhibitor (SARI) (Fagiolini et al., 2012) that might also contribute to the observed effects. Potentially, SSRIs and SARIs that increase extracellular serotonin by inhibiting its uptake, can then attenuate effects of psychedelics via receptor competition with the endogenous serotonin. In addition, it has been shown that repeated administration of SSRIs desensitises 5-HT_2A_ receptors which may reduce the cell’s response to psychedelics by binding to these receptors (Gray & Roth, 2001). Chronic SSRI use can also increase serotonin release via desensitisation of 5-HT_1A_ raphe autoreceptors (Artigas et al., 2001).

However, one SSRI **escitalopram,** did not have an effect on psilocybin-induced positive mood or mind-altering effects (depersonalisation, oceanic boundlessness or euphoria), but it did reduce ego disintegration and anxiety (Becker et al., 2022). The authors hypothesised that while LSD does not affect the serotonin transporter, psilocin has a weak inhibitory effect on it (Rickli et al., 2016). This distinction in pharmacology could lead to different interactions between antidepressants and LSD compared to psilocybin (Becker et al., 2022), explaining the differences in response. Although this study offers important insights into the safety of administering psilocybin to patients taking escitalopram, it is crucial to bear in mind that the treatment phase of this study was 14 days. Longer treatment periods with escitalopram, which may often last for years, could lead to distinct outcomes due to molecular changes that occur over extended periods (Faure et al., 2006).

SSRIs can also result in DDI at the pharmacokinetic level as some of them, such as fluoxetine, are potent inhibitors of CYP2D6 enzymes (Brøsen, 1998), which can modulate the effects of psychedelics that are being metabolised by such enzymes. For example, CYP2D6 enzymatic activity is already known to influence the effects of LSD (Vizeli et al., 2021). However, in the case of a reduced CYP2D6 activity, a stronger and longer response would have been expected for LSD when it was used with fluoxetine, sertraline or paroxetine. Ultimately, this can be an interplay between different molecular actions, including interactions with enzymes, receptor availability and other molecular factors.

Results from another antidepressant class, MAOIs, showed a similar outcome to those of SSRIs. In particular, **phenelzine**, **isocarboxazid**, **nialamide** and **iproniazid** are all non-selective MAO inhibitors (Entzeroth & Ratty, 2017), therefore inhibiting both MAO-A and MAO-B. The first three listed antidepressants attenuated or blocked the effects of LSD (Bonson & Murphy, 1996; Grof & Dytrych, 1965; Resnick et al., 1964). In contrast, the latter, iproniazid, did not alter the subjective or physical effects of LSD (DeMaar et al., 1960). However, iproniazid was also used in the experiments with DMT where it was shown to reduce the effects of DMT (Sai Halasz, 1963). This particular finding is interesting as iproniazid is an irreversible MAOI (Entzeroth & Ratty, 2017) which inhibits the enzyme that rapidly metabolises DMT (Barker et al., 1980). Moreover, studies in rats have shown that iproniazid pretreatment increases levels of DMT in the brain, as well as in the liver, kidney and blood (Sitaram et al., 1987). However, the reduced DMT effect could be due to increased serotonin levels after blocking MAO and it is hypothesised that higher doses of DMT are needed when serotonin levels are elevated (Sai Halasz, 1963). It is suggested that ayahuasca’s effect is mediated by MAO inhibition in the digestive system or bloodstream, which protects DMT from metabolism during its journey to the brain, where MAO inhibitors can then attenuate DMT’s effects due to elevated brain serotonin (Ott, 1996). This hypothesis for DMT can also explain the similar, attenuated response which was observed in the studies where LSD was combined with MAOIs.

In addition to metabolising DMT, MAOIs are also important in the breakdown of serotonin as they are blocking MAO enzymes involved in its metabolism (Foong et al., 2018). This gives rise to a possible scenario of excessive serotonin levels in the brain. In particular, when combining MAOIs with each other or with SSRI or SNRI, it has been thought to carry the greatest risk of serotonin toxicity (Foong et al., 2018). In one case report, symptoms resembling serotonin toxicity were reported after an individual administered ayahuasca while being treated on a **fluoxetine** treatment (Callaway & Grob, 1998). However, the patient also recovered rapidly within four hours after the administration of ayahuasca without any treatment. Moreover, another case report that was not included in the review, documented an instance of serotonin toxicity in an individual who had been undergoing **fluoxetine and quetiapine** treatment while consuming pure harmal extracted from *Peganum harmala*, which is a component of ayahuasca (Bakim et al., 2012).

Ayahuasca brew is usually made from a plant containing DMT and another plant that contains β-carbolines which mostly inhibit MAO-A, thereby making DMT orally active (McKenna, 2004). Tetrahydroharmine, one of the β-carbolines present in the brew, can also act as a weak serotonin uptake inhibitor and increase brain serotonin levels (Airaksinen et al., 1980). Fluoxetine strongly inhibits the CYP2D6 enzyme, which metabolises many drugs, including serotonin. The concurrent inhibition of serotonin reuptake and serotonin-metabolising enzymes, such as CYP2D6 and MAO, can cause an accumulation of serotonin in the brain and low clearance, potentially leading to life-threatening serotonin toxicity (Dunkley et al., 2003). This explanation aligns with the reported case reports; however, further research is necessary to confirm the potential association.

One study that reported a blocking action on LSD effects was using a high dose of **niacin** (vitamin B3). Pretreatment with niacin resulted in delayed onset of LSD-effects and prevented most of the perceptual changes from occurring, and administration post-LSD ingestion attenuated all effects of LSD within 5 minutes (Agnew & Hoffer, 1955). The blocking mechanism of niacin is not clear, but may be due to increased serotonin levels. This is indicated by the studies where niacin has been shown to induce serotonin release from human platelets within a few minutes and to increase plasma serotonin levels in rats (Papaliodis et al., 2008). Secondly, blocking 5-HT_2A_ receptors with ketanserin inhibits niacin-induced temperature increase (Papaliodis et al., 2008), which indicates that serotonin is involved in the effects of niacin. Finally, high-dose treatment with niacin has caused manic-like psychotic episodes that were thought to occur via stimulation of serotonin, but also dopamine production (Loebl & Raskin, 2013).

Several studies have reported the potentiated effects of LSD. In one study, **desipramine**, **imipramine** and **clomipramine,** which are all tricyclic antidepressants, were shown to increase psychological effects of LSD (Bonson & Murphy, 1996). Desipramine is a potent inhibitor of noradrenaline reuptake, while imipramine and clomipramine exhibit a lower degree of noradrenaline and serotonin reuptake inhibition (Gillman, 2007). Furthermore, clomipramine also functions as a 5-HT_2A_ receptor antagonist and also imipramine has affinity for this receptor subtype (Gillman, 2007).

Since TCAs were reported to enhance the effects of LSD despite some of them inhibiting serotonin uptake and are even 5-HT_2A_ receptor antagonists (similar to SSRIs that decreased the effects of LSD), it is suggested another mechanism can contribute. For instance, chronic administration of TCAs, such as desipramine, can increase the sensitivity of certain neurons to LSD, suggesting that these medications may sensitise postsynaptic serotonin receptors in the brain and therefore be more responsive to LSD (Bonson & Murphy, 1996). Alternatively, the effects can be enhanced through a dopaminergic system (Bonson & Murphy, 1996) as chronic use of desipramine has been demonstrated to result in an elevated behavioural response to amphetamine, whereas chronic use of fluoxetine did not have the same effect (Spyraki & Fibiger, 1981). Therefore, it is possible that the observed effects are due to a modification in the sensitivity of the dopamine receptors.

Additionally, from the same study, it was reported that chronic **lithium** use potentiated LSD effects and resulted in earlier onset (Bonson & Murphy, 1996). It has been suggested that while acute administration of lithium increases serotonin levels in the brain, chronic administration on the other hand reduces serotonin concentrations (Bonson & Murphy, 1996). This may explain why long-term lithium use enhances LSD effects, as LSD acts as an agonist in the absence of endogenous serotonin resulting in the observed behavioural effects (Bonson & Murphy, 1996). It is important to note that these findings and some anecdotal reports that can be found on internet forums suggesting a similar outcome, there are also several reported instances of lithium causing seizures when used concurrently with LSD or psilocybin (Nayak et al., 2021).

Intensified and prolonged (negative) effects of LSD have also been reported in individuals being pretreated with **reserpine**, an antihypertensive drug, in three separate studies (Freedman, 1963; Isbell & Logan, 1957; Resnick et al., 1965). Freedman (1963), and Isbell and Logan (1957) both reported tremors that were present after using LSD together with reserpine. Tremors were not observed during LSD use alone (Isbell & Logan, 1957), and when combined with reserpine, the experience was reported to be less pleasant and lasted longer than LSD use alone (Freedman, 1963). Furthermore, specific types of hallucinations were reported with reserpine treatment that were not observed during LSD use alone. Overall, individuals reported unpleasant experiences when using an LSD with reserpine.

Reserpine has the ability to bind to the storage vesicles of certain neurotransmitters, including dopamine and norepinephrine. This binding inhibits the catecholamine pumps and blocks the uptake of serotonin, norepinephrine and dopamine into the presynaptic storage vesicles. Ultimately, this leads to the depletion of these neurotransmitters by cytoplasmic MAO at both central and peripheral synapses (Cheung & Parmar, 2023). Lower serotonin levels can enhance LSD effects, which was also observed for lithium. However, when LSD was used after chronic-administration of lithium, it resulted in positive effects and more vivid hallucinations, without any noted increase in negative psychological or physical side effects as for reserpine. Treatment with reserpine has been known to cause several neurological side effects (Cheung & Parmar, 2023) and studies have shown that in patients with anxiety it can exacerbate their symptoms (Peterfy et al., 1976; Sarwer-Foner & Ogle, 1956). Since LSD can also increase anxiety as a side effect (Strassman, 1984), it is possible that reserpine treatment could enhance this effect (or *vice versa*), while the modulated hallucinogenic effects of LSD are due to depleted serotonin levels.

Moreover, reserpine is also a potent inhibitor of both CYP2C19 and CYP2D6 enzymes (Englund et al., 2014), which are involved in the initial metabolic steps of LSD (Wagmann et al., 2019). This inhibition may account for the reported prolonged effects of LSD when it was used with reserpine (Freedman, 1963) as it could potentially decrease the metabolism of LSD. Finally, reserpine acts as an inhibitor of P-glycoprotein as well (Englund et al., 2014), which can lead to increased concentrations of P-gp substrates. However, no research has been conducted to investigate the impact of P-gp activity on LSD pharmacokinetics and its effects, making it difficult to assess the potential contribution of P-gp inhibition.

A 5-HT_1_ receptor antagonist, **racemic pindolol** was reported to intensify the effects of DMT (Strassman, 1996). Pindolol is a β-adrenoceptor and 5-HT_1A_ antagonist (high affinity) (Artigas et al., 2001). The authors suggested a buffering effect of 5-HT_1A_ that blocked the 5-HT_2_ mediated psychedelic effects (Strassman, 1996). However, pindolol has been shown to accelerate, and in some cases, enhance the antidepressant effects of SSRIs. This can be mediated by antagonising 5-HT_1A_ autoreceptors in the midbrain raphe and, as a result, preventing the inhibition of serotonin release (Artigas et al., 2001). DMT is a full agonist at 5-HT_1A_ receptors and has higher affinity for the receptor than it has for 5-HT_2A_ receptors where it is weak partial agonist (Kozell et al., 2023). Hypothetically, a similar effect as for SSRIs has been described, can also modulate the effects of DMT, which may have enhanced effects via activating postsynaptic 5-HT_1A_ receptors while 5-HT_1A_ autoreceptors are blocked.

A 5-HT_1_ receptor agonist and 5-HT_2_ receptor antagonist **methysergide** was similarly shown to potentiate DMT’s effects (Sai-Halasz, 1962). This finding adds more weight on the 5-HT_1A_ receptor contribution to the effects of DMT, especially considering the methysergide also an antagonist at the 5-HT_2A_ receptors.

With regards to mescaline use and potentiated effects, one case report involved the use of **2C-O**, a structural isomer of mescaline that did not appear to have psychedelic properties on its own (Dittrich, 1971). However, when used together with mescaline, a synergistic effect may have occurred that potentiated the effects of mescaline described in the case report.

**Azacyclonol**, once investigated as a potential antidepressant, was experimented in individuals who administered LSD and mescaline. Two studies showed no effect of blocking LSD’s actions (Clark, 1956; Isbell & Logan, 1957) and one case report describes the same for mescaline (Clark, 1956). These results are in contrast to another study that reported the attenuated (pretreatment) or blocked (acute) effects of mescaline after azacyclonol treatment (Fabing, 1955). The mechanism of action of azacyclonol is not well understood, and the mixed results from the described studies make it difficult to draw firm conclusions and hypotheses.

Finally, two studies reported the effects of recreational drugs, such as alcohol and cannabis consumption on LSD or psilocybin. In one case report, **cannabis** induced hallucinogen persisting perception disorder (Espiard et al., 2005). The second study, which investigated the effects of **alcohol**, found that LSD and psilocybin (to a lesser extent) acted as antagonists to the subjective effects of alcohol, while their own psychedelic effects were mainly unaffected (Barrett et al., 2000). It has been suggested that LSD may block the subjective effects of alcohol by interacting with 5HT_1B_ and 5HT_1C_ receptors, which are implicated in the formation of the ethanol cue (Barrett et al., 2000). Evidence shows that agonists of these receptors produce responses similar to those of alcohol, whereas blockade of these receptors interferes with the discriminative stimulus properties of alcohol (Barrett et al., 2000).

### Limitations

One of the limitations of this study is the inclusion of a number of old research articles, particularly those published between the 1950s and the 1970s, where many of them provided limited information about the outcomes and/or methods used. Additionally, the limited number of total studies included in this review led to the inclusion of case reports, which may be subject to bias and may provide limited generalizability to larger populations. This review may also have also missed some relevant studies that were published only in non-English languages, which were more common in the early days of research.

## Conclusions

In this systematic review, we observed DDIs at both pharmacodynamic and (likely) pharmacokinetic levels that may block or decrease the response to psychedelics, or alternatively potentiate and lengthen the duration of psychological and/or physical effects. While there is strong evidence of 5-HT_2A_ receptor involvement in the effects of psychedelics, some research included in this review suggests that other serotonin receptors, such as 5-HT_1A/B_ and dopamine receptors, along with altered serotonin levels, may also modulate psychological and/or physical effects. Additionally, a small number of studies reviewed indicated a potential role of the 5-HT_1_ receptor subtype in modulating effects of DMT. It appears that although different psychedelics may yield similar subjective effects, their pharmacological properties differ, resulting in potentially varying interaction effects when combined with other psychoactive drugs. Overall, given the limited number of papers exploring DDIs associated with psychedelics and the resurgence of scientific and medical interest in these compounds, further research is needed to improve understanding of such interactions, and identify novel drug interactions and potentially serious adverse reactions not currently described in the literature.

## Data Availability

All data produced in the present work are contained in the manuscript.

## Acknowledgments

The authors acknowledge Eleanor White for her contribution to an earlier version of the introduction.

