## Supplementary Information for "Drug-drug interactions between classic psychedelics and psychoactive drugs: a systematic review"

### Search terms

**The following search was used in PubMed:**
(

"Lysergic Acid Diethylamide"[MeSH] OR

"lysergic acid diethylamide*"[Title/Abstract] OR

"LSD"[Title/Abstract] OR

"L.S.D."[Title/Abstract] OR

"Psilocybin"[MeSH] OR

"psilocybin"[Title/Abstract] OR

"psilocin"[Title/Abstract] OR

"magic mushroom*"[Title/Abstract] OR

"Psilocybe"[Title/Abstract] OR

"Mescaline"[MeSH] OR

"mescalin*"[Title/Abstract] OR

"peyote"[Title/Abstract] OR

"san pedro"[Title/Abstract] OR

"3,4,5-trimethoxyphenethylamine"[Title/Abstract] OR

"trimethoxyphenethylamine"[Title/Abstract] OR

"N,N-Dimethyltryptamine"[MeSH] OR

"dimethyltryptamine"[Title/Abstract] OR

"DMT"[Title/Abstract] OR

"D.M.T."[Title/Abstract] OR

"ayahuasca"[Title/Abstract] OR

"harmaline"[Title/Abstract] OR

"harmine"[Title/Abstract] OR

"banisteriopsis"[Title/Abstract] OR

"psychotria"[Title/Abstract] OR

"chacruna"[Title/Abstract] OR

"diplopterys"[Title/Abstract] OR

"chaliponga"[Title/Abstract] OR

"chagropanga"[Title/Abstract] OR

"syrian rue"[Title/Abstract] OR

"peganum"[Title/Abstract] OR

"mimosa"[Title/Abstract] OR

"jurema"[Title/Abstract] OR

"acacia"[Title/Abstract]

)

AND

(

"Drug Interactions"[MeSH] OR

"Drug-Related Side Effects and Adverse Reactions"[MeSH] OR

"Pharmacokinetics"[MeSH] OR

"drug-drug"[Text Word] OR

"co-administrat*"[Text Word] OR

"co-ingest*"[Text Word] OR

"interaction"[Text Word] OR

"pharmacodynamic*"[Text Word] OR

"pharmacokinetic*"[Text Word]

)

NOT

(

"rat"[Title] OR

"rats"[Title] OR

"mouse"[Title] OR

"mice"[Title] OR

"murine"[Title] OR

"cat"[Title] OR

"cats"[Title] OR

"dog"[Title] OR

"dogs"[Title]

)

AND

(

"journal article"[Publication Type] OR

"randomized controlled trial"[Publication Type] OR

"clinical study"[Publication Type] OR

"clinical trial"[Publication Type] OR

"clinical trial, phase i"[Publication Type] OR

"clinical trial, phase ii"[Publication Type] OR

"clinical trial, phase iii"[Publication Type] OR

"clinical trial, phase iv"[Publication Type] OR

"controlled clinical trial"[Publication Type] OR

"pragmatic clinical trial"[Publication Type] OR

"comparative study"[Publication Type] OR

"observational study"[Publication Type] OR

"case reports"[Publication Type]

)

NOT

(

"review"[Publication Type] OR

"preprint"[Publication Type]

)

AND

(

"english"[Language]

)

**The following search was used in Web of Science:**

TS=("lysergic acid diethylamide*" OR LSD OR "L.S.D." OR psilocybin OR psilocin OR "magic mushroom*" OR Psilocybe OR mescalin* OR peyote OR "san pedro" OR "3,4,5-trimethoxyphenethylamine" OR "trimethoxyphenethylamine" OR "N,N-Dimethyltryptamine" OR "dimethyltryptamine" OR "DMT" OR "D.M.T" OR harmaline OR harmine OR ayahuasca OR banisteriopsis OR psychotria OR chacruna OR diplopterys OR chaliponga OR chagropanga OR "syrian rue" OR peganum OR mimosa OR jurema OR acacia)

AND ALL=(drug-drug OR co-administrat* OR co-ingest* OR interaction OR pharmacodynamic* OR pharmacokinetic*)

NOT TI=(rat OR rats OR mouse OR mice OR murine OR cat OR cats OR dog OR dogs)

AND DT=(Article)

AND LA=(English)

AND SU=(Pharmacology OR Pharmacy OR Neurosciences OR Psychiatry OR Clinical Neurology OR Biochemistry Molecular Biology OR Chemistry Medicinal OR Psychology Clinical OR Toxicology OR Multidisciplinary Sciences OR Psychology OR Substance Abuse OR Medicine Research Experimental OR Behavioral Sciences OR Biology OR Integrative Complementary Medicine OR Cell Biology OR Medicine General Internal OR Psychology Biological OR Public Environmental Occupational Health OR Medicine Legal OR Physiology OR Immunology OR Psychology Experimental OR Pediatrics OR Emergency Medicine OR Medical Ethics OR Genetics Heredity)

**The following search was used in PsycINFO:**

(

exp "lysergic acid diethylamide"/ or

"lysergic acid diethylamide*".ti,ab. or

"LSD".ti,ab. or

"L.S.D.".ti,ab. or

exp "psilocybin"/ or

"psilocybin".ti,ab. or

"psilocin".ti,ab. or

"magic mushroom*".ti,ab. or

"psilocybe".ti,ab. or

exp "mescaline"/ or

"mescalin*".ti,ab. or

"peyote".ti,ab. or

"san pedro".ti,ab. or

"3,4,5-trimethoxyphenethylamine".ti,ab. or

"trimethoxyphenethylamine".ti,ab. or

exp "n,n-dimethyltryptamine"/ or

"dimethyltryptamine".ti,ab. or

"dmt".ti,ab. or

"d.m.t.".ti,ab. or

"ayahuasca".ti,ab. or

"harmaline".ti,ab. or

"harmine".ti,ab. or

"banisteriopsis".ti,ab. or

"psychotria".ti,ab. or

"chacruna".ti,ab. or

"diplopterys".ti,ab. or

"chaliponga".ti,ab. or

"chagropanga".ti,ab. or

"syrian rue".ti,ab. or

"peganum".ti,ab. or

"mimosa".ti,ab. or

"jurema".ti,ab. or

"acacia".ti,ab.

)

and

(

exp "drug interactions"/ or

exp "drug-related side effects and exp adverse reactions"/ or

exp "pharmacokinetics"/ or

"drug-drug".tw. or

"co-administrat*".tw. or

"co-ingest*".tw. or

"interaction".tw. or

"pharmacodynamic*".tw. or

"pharmacokinetic*".tw.

)

not

(

"rat".ti or

"rats".ti or

"mouse".ti or

"mice".ti or

"murine".ti or

"cat".ti or

"cats".ti or

"dog".ti or

"dogs".ti

)

* Search results limited to: Journal article or Reprint (Document type); Humans (Population group); English (Language)
